# High-resolution mapping and characterization of epitopes in COVID-19 patients

**DOI:** 10.1101/2020.11.23.20235002

**Authors:** Winston A. Haynes, Kathy Kamath, Joel Bozekowski, Elisabeth Baum-Jones, Melissa Campbell, Arnau Casanovas-Massana, Patrick S. Daugherty, Charles S. Dela Cruz, Abhilash Dhal, Shelli F. Farhadian, Lynn Fitzgibbons, John Fournier, Michael Jhatro, Gregory Jordan, Debra Kessler, Jon Klein, Carolina Lucas, Larry L. Luchsinger, Brian Martinez, Mary C. Muenker, Lauren Pischel, Jack Reifert, Jaymie R. Sawyer, Rebecca Waitz, Elsio A. Wunder, Minlu Zhang, Yale IMPACT Team, Akiko Iwasaki, Albert I. Ko, John C. Shon

## Abstract

Fine scale delineation of epitopes recognized by the antibody response to SARS-CoV-2 infection will be critical to understanding disease heterogeneity and informing development of safe and effective vaccines and therapeutics. The Serum Epitope Repertoire Analysis (SERA) platform leverages a high diversity random bacterial display library to identify epitope binding specificities with single amino acid resolution. We applied SERA broadly, across human, viral and viral strain proteomes in multiple cohorts with a wide range of outcomes from SARS-CoV-2 infection. We identify dominant epitope motifs and profiles which effectively classify COVID-19, distinguish mild from severe disease, and relate to neutralization activity. We identify a repertoire of epitopes shared by SARS-CoV-2 and endemic human coronaviruses and determine that a region of amino acid sequence identity shared by the SARS-CoV-2 furin cleavage site and the host protein ENaC-alpha is a potential cross-reactive epitope. Finally, we observe decreased epitope signal for mutant strains which points to reduced antibody response to mutant SARS-CoV-2. Together, these findings indicate that SERA enables high resolution of antibody epitopes that can inform data-driven design and target selection for COVID-19 diagnostics, therapeutics and vaccines.

## Introduction

The novel coronavirus SARS-CoV-2 global pandemic has affected millions of people world-wide and led to a major healthcare crisis. Considerable research has gone into understanding the myriad symptoms that are seen in patients as well as the stark contrast between the large number of mild or asymptomatic cases and the staggering death toll around the world^1–5^. Determining the factors that contribute to different disease manifestations, severity and immunity is critical to adequate therapeutic intervention, improved patient outcomes, and vaccine design.

One avenue that is being extensively explored is the degree to which an immune response to the virus protects, or harms, an individual. Although it is possible that a pre-existing exposure to common coronaviruses may have a protective role during SARS-CoV-2 infection^6,7^, it has also been proposed that antibodies to SARS-CoV-2 may sometimes be directly pathogenic or lead to the generation of auto-reactive antibodies^8–12^. With millions of cases extant, and based on current trends, millions more in the coming months, it is critical that patients be accurately assessed not just for infection but also for the potential of severe disease progression, allowing timely application of treatments for best outcomes. Of considerable concern as well is the specter of a combined SARS-CoV-2/influenza season with the need to rapidly differentiate between multiple viral infections^13,14^. In addition, a growing number of COVID-19 patients who had expected to fully recover have not, with symptoms that linger far past the expected recovery period and cause significant disruption to their lives as well as an extended need for healthcare. The number of “long-haulers” is not currently clear but the need to elucidate the role of a disrupted immune system in their illness is pressing^15–17^.

Of paramount urgency is the development of a vaccine against SAR-CoV-2. Along with the initial step of defining an effective vaccine for the immediate crisis, factors such as viral mutation rate and the uncertainty of long-term immunity could play a large role in ongoing management. It is unclear if it will be possible to develop “sterilizing immunity” to the virus, thus preventing infection completely^18–20^. A yearly “flu-type” immunization would necessitate continued surveillance of both viral evolution and patients’ yearly immune responses to keep transmission and mortality to a minimum^21^.

Many different groups have examined antibody responses to SARS-CoV-2, exploring correlation with disease severity, duration of humoral response, and the neutralizing capacity of response^3,22,23^. Most of these methods have been limited to quantitative assessment of humoral response to whole proteins or large domains of spike and nucleoprotein. Peptide and phage display libraries have also been used to capture higher resolution epitope patterns associated with disease but are limited to characterization of linear epitope signal and in their ability to make clinical seropositivity assessments^4,24,25^. We present in this paper the application of Serum Epitope Repertoire Analysis (SERA), a high throughput, random bacterial peptide display technology that enables assessment of SARS-CoV-2 seropositivity and high-resolution mapping of epitopes across any arbitrary proteome, including wild-type SARS-CoV-2, its mutant strains, common coronaviruses, and the human proteome.

We have leveraged over 2,000 pre-pandemic immune repertoires and over 500 COVID-19 cases to identify the antigens and epitopes that elicit a SARS-CoV-2 humoral response. We show that while antibody profiles of individuals are heterogeneous, epitope-level resolution enables a range of analyses and visualizations, from the earliest epitopes to elicit an antibody response, to putative mapping of structural epitopes that may be important for neutralization or immunity. Combining epitope motifs into a panel yields a diagnostic classifier that distinguished NAT+ cases from controls with accuracy comparable to serological tests in current use. Differences in the quantity and quality of epitopes in mild versus moderate and severe disease can be seen at sites of biological and clinical interest. *In silico* analysis of epitope repertoires on wild-type and mutant SARS-CoV-2 proteins suggests that some mutations may result in loss of antibody reactivity to mutant SARS-CoV-2 infections while analysis against the human proteome identified SARS-CoV-2 antibodies that may cross-react with human proteins and contribute to disease pathogenesis. These capabilities are all possible through informatics analysis of a single assay that requires a minimal amount of serum from each subject.

## Results

### SERA screening of COVID-19 serum

We applied SERA to discover and validate SARS-CoV-2 antigens and epitopes across the complete viral proteome from 779 COVID-19 serum samples taken from multiple cohorts of individuals with recent or past SARS-CoV-2 infection, which in total include 579 unique subjects (Table 1). We additionally leverage a large database of 1997 pre-pandemic controls. The majority of the subjects were confirmed SARS-CoV-2 positive by nucleic acid testing (NAT). For Cohorts I, II, and III, extensive characterization was available for covariates that included disease severity, date of symptom onset, and in many cases, serological testing (Supplemental Table 1).

**Table 1:** SARS-CoV-2 cohorts used for epitope motif discovery.

| SARS-CoV-2 Cohorts |  |  | Training |  | Testing |  |  |
| --- | --- | --- | --- | --- | --- | --- | --- |
|  |  |  | # of Donors | # of Samples | # of Donors | # of Samples | COVID-19 Test |
| Cohort I | Yale | Inpatient | 91 | 153 | 98 | 177 | NAT and/or serology |
|  |  | Healthcare Workers | 4 | 4 | 6 | 9 | NAT and/or serology |
|  |  | Outpatient | 4 | 4 | 5 | 5 | Serology or symptoms |
| Cohort II | LabCorp | Inpatient |  |  | 188 | 235 | NAT |
|  |  | Outpatient |  |  | 10 | 10 | Serology |
| Cohort III | SBCH | In and Outpatient |  |  | 73 | 82 | NAT, Serology |
| Cohort IV | BioIVT | In and Outpatient |  |  | 21 | 21 | Serology (20), NAT (1) |
| Cohort V | BCA | Asymptomatic/Mild |  |  | 79 | 79 | Serology/SERA |
|  | Total |  | 99 | 161 | 480 | 618 |  |
|  | Prepandemic Controls | IgG | 497 | 497 | 1500 | 1500 |  |
|  |  | IgM | 430 | 430 | 1498 | 1498 |  |

Patient samples were all screened using the previously published SERA assay, which enables high throughput characterization of antibody epitopes (Figure 1)^26,27^. In brief, serum or plasma is incubated with the randomized bacterial display peptide library; antibodies bind to peptides that mimic their natural epitopes and are then separated from unbound library members using affinity-coupled magnetic beads. The resulting bacterial pools are grown overnight, plasmids encoding the antibody-binding peptides are purified, and the peptide-encoding regions are PCR amplified and barcoded with well-specific PCR indices. Ninety-four samples are normalized, pooled together and sequenced via next-generation sequencing (NGS). The output of SERA is a set of approximately 1 million peptide sequences for each individual, representing their unique epitope repertoire. After SERA screening, we applied two complementary discovery tools, IMUNE and PIWAS, to identify antigens and epitopes involved in the SARS-CoV-2 immune response (Figure 1).

**Figure 1:**
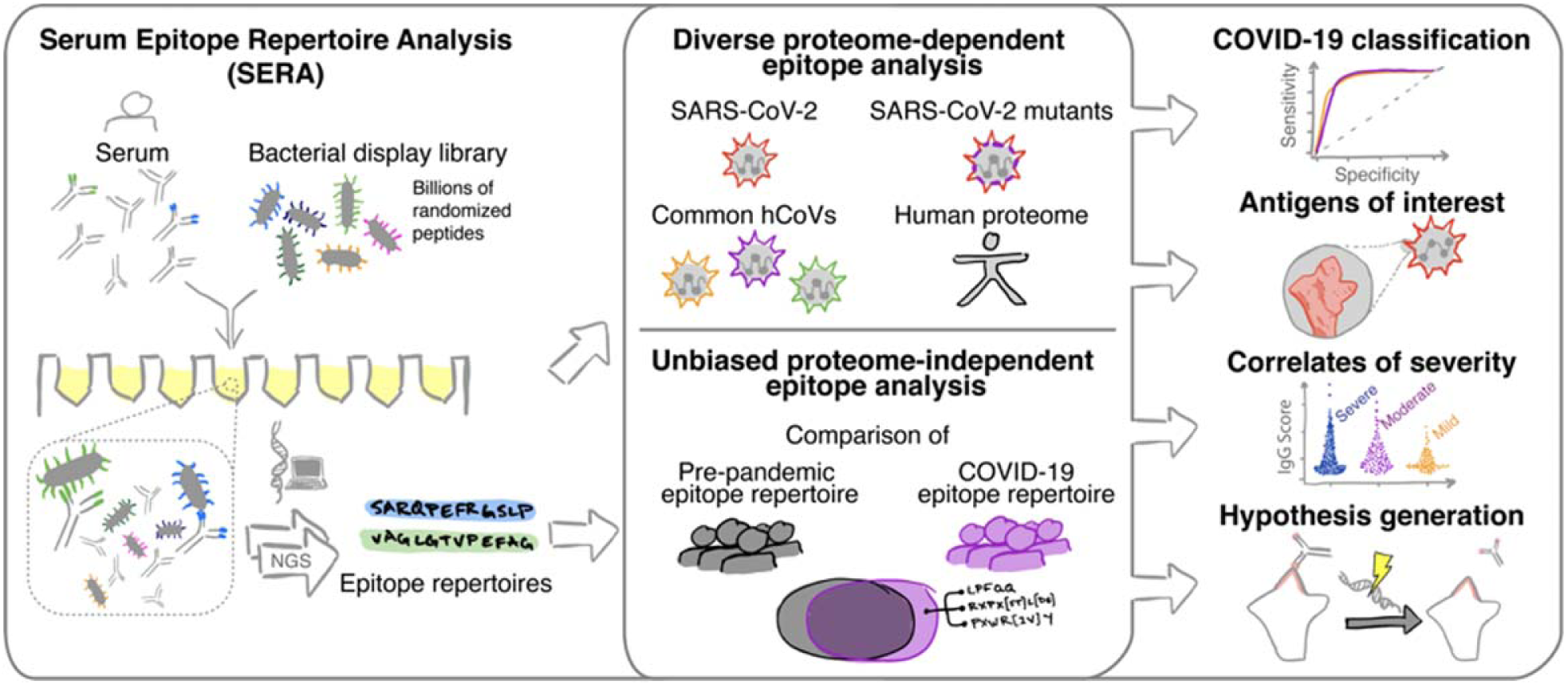
The Serum Epitope Repertoire Analysis (SERA) platform enables high resolution mapping of SARS-CoV-2 antibody repertoires. The SERA assay results in a set of approximately 1 million unique peptides, the “epitope repertoire”, for each individual. Repertoires were deposited in a database and compared with pre-pandemic controls to identify conserved epitopes in SARS-CoV-2 using proteome-dependent and -independent bioinformatic methods. SERA enables analysis of COVID-19 repertoires against any proteome including mutant SARS-CoV-2 strains, human common coronaviruses and the human proteome for discovery of potential autoantigens. The identified epitope signatures can be used to build diagnostic classifiers, to identify correlates of disease severity, and to develop hypotheses based on cases with specific symptoms and/or disease course (neurological, GI, cardio e.g.).

### Characterization of SARS-CoV-2 proteome antigens and epitopes

To establish an understanding of relevant SARS-CoV-2 antigens and epitopes, we analyzed the SARS-CoV-2 proteome with protein-based immunome wide association studies (PIWAS). Briefly, PIWAS identifies epitope signal in the context of an arbitrary proteome by tiling and smoothing kmer sequences across the entire proteome^28^. PIWAS derives power at both the cohort and single sample level through statistical comparisons to a large database of pre-pandemic controls. Using the reference SARS-CoV-2 proteome from Uniprot, we performed PIWAS of 579 COVID-19 samples compared to 497 pre-pandemic controls, with 1500 additional pre-pandemic controls serving as a normalization cohort. In addition to the established antigens spike and nucleocapsid, we observed highly significant signals for protein 3a, non-structural protein 8 (NSP-8), membrane protein, and replicase polyprotein 1ab (Figure 2A). We further examined epitope-level signal for the top IgG and IgM antigens identified by PIWAS (Figure 2B). Within spike and nucleoprotein, we observed multiple epitopes that are conserved across a large portion of the COVID-19 patient population. In contrast, epitope signals for protein 3a, NSP-8, and membrane protein (IgM) are largely characterized by a single, dominant epitope. While the receptor binding domain (RBD) of spike is highly important in host infection by the virus, we observe no conserved epitope signal against this region of spike (amino acids 331-524). Instead, we observe private spike epitopes in a subset of patients in our cohorts (Figure 2C, Figure S1). We highlight patients with epitopes observe in multiple longitudinal draws, to decrease the likelihood of false positive signal.

**Figure 2:**
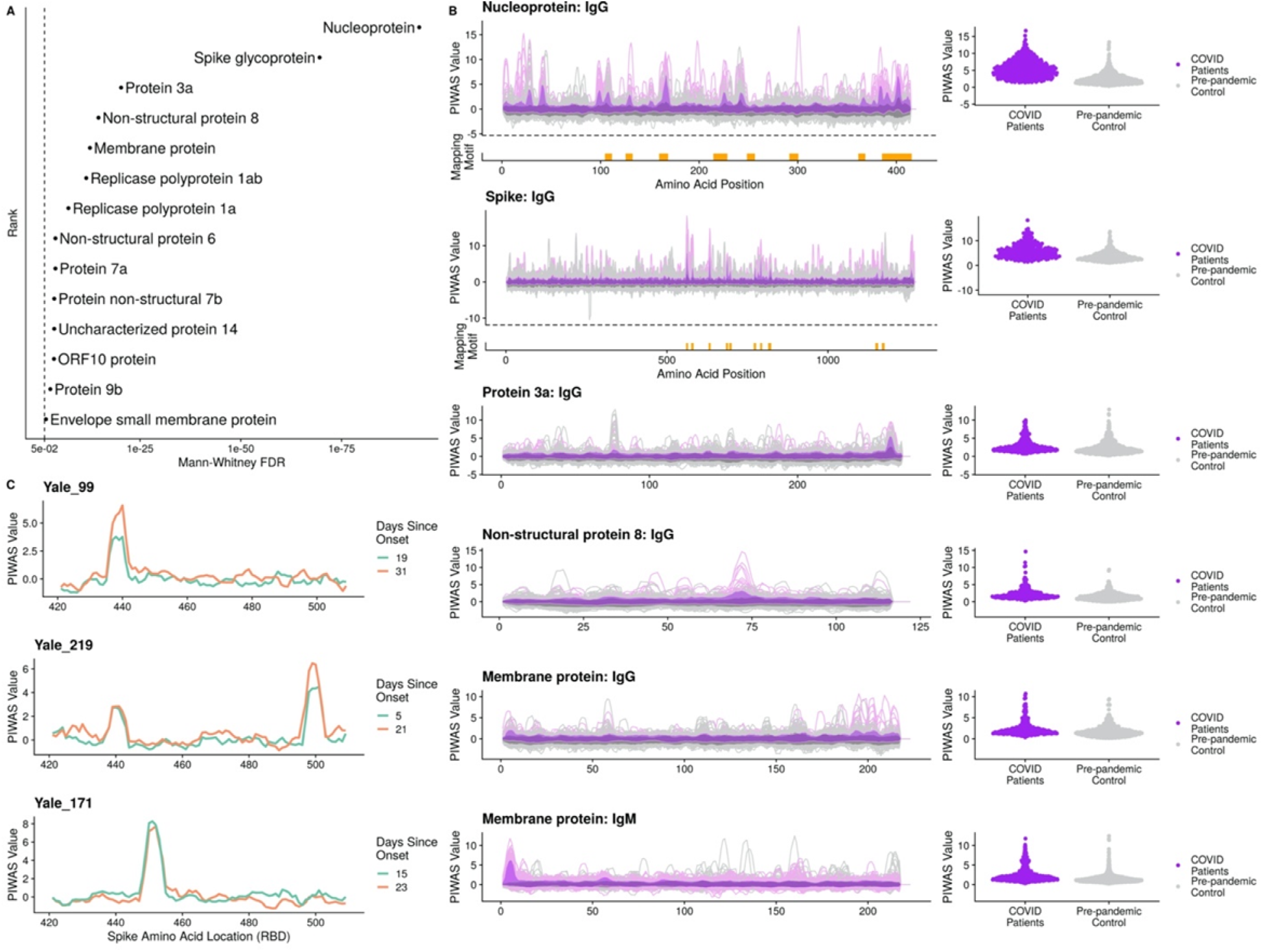
Bioinformatic analysis of SERA antibody repertoires identifies the antigens and epitopes involved in the SARS-CoV-2 immune response. (**A**) PIWAS statistical ranking of kmer enrichments across the SARS-CoV-2 proteome using the Mann-Whitney false discovery rate (FDR). Multiple antigens in addition to spike and nucleoprotein showed significant enrichment for one or more epitopes. (**B**) PIWAS kmer enrichments from COVID-19 repertoires versus pre-pandemic controls across statistically significant antigens. PIWAS values = number of standard deviations above the mean of 1500 pre-pandemic controls. IMUNE motifs largely mapped to the same epitopes that were identified by PIWAS. Epitopes on spike and nucleoprotein discovered by IMUNE are shown below each antigen (orange bars). (**C**) Longitudinal samples from individual subjects enabled identification of RBD-specific signals that emerged over time but were not conserved across COVID-19 patients.

### Unbiased, proteome-independent epitope analysis

The IMUNE algorithm identified mapping and non-mapping epitope motifs that were highly enriched in COVID-19 repertoires (Methods)^27^. Linear epitopes identified by IMUNE largely overlapped with those identified by PIWAS (Figure 2). The IgG linear motifs mapped to epitopes on spike glycoprotein (n=10), nucleoprotein (n=8) and NSP8 (n=2). IgM linear motifs mapped to a single epitope at the furin cleavage site on spike glycoprotein that was also a target for IgG antibodies, as well as one epitope on the SARS-CoV-2 membrane protein. A significant number of motifs identified by IMUNE did not directly map to linear regions of the SARS-CoV-2 proteome. We have observed from studies with monoclonal antibodies that non-mapping motifs tend to represent mimotopes of both linear and structural epitopes.

Motifs were selected for inclusion in the SARS-CoV-2 epitope map if they demonstrated a specificity of at least 98% in 497 pre-pandemic controls (Methods). The resulting SARS-CoV-2 panel of 45 IgG and 14 IgM motifs was compiled into a semi-quantitative epitope map, enabling visualization of motif enrichment for all evaluated COVID-19 and control samples (Figure 3A). We observed that an unlabeled, hierarchical clustering of samples based on these motif enrichments largely separates pre-pandemic control samples from COVID-19 patients. Focusing on those motifs with linear hits to SARS-CoV-2, we further observed sub-clusters of patients with reactivity to specific isotypes and antigens, from left-to-right: spike IgG+IgM, spike and membrane IgM, spike IgM, nucleocapsid IgG, and broadly reactive.

**Figure 3:**
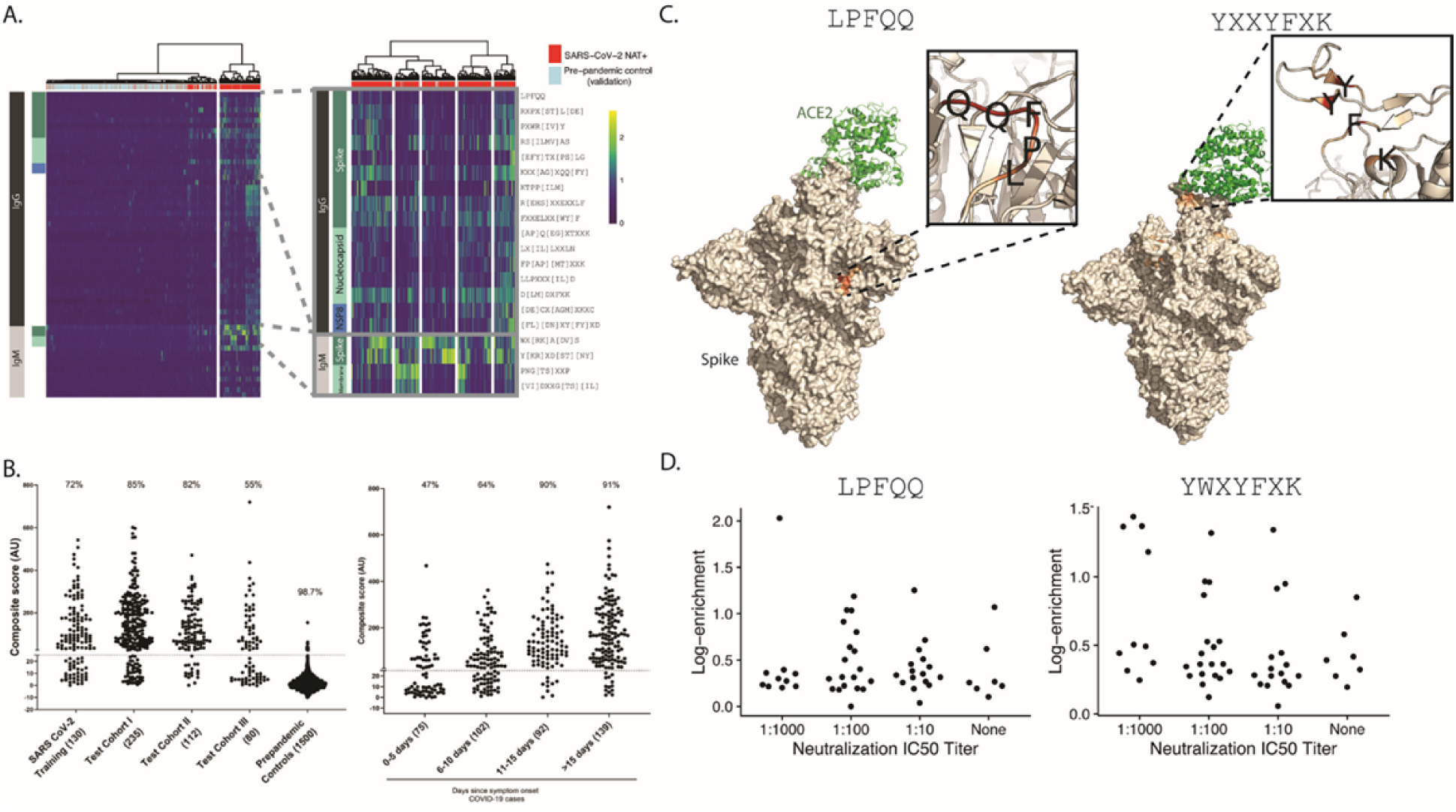
IMUNE-based discovery of IgG and IgM motifs in the SARS-CoV-2 humoral immune response. (**A**) Heatmap of IgG and IgM motif log-enrichment values for 579 COVID-19 samples and 1500 pre-pandemic controls. Inset highlights motifs with linear epitope maps to SARS-CoV-2. (**B**) Sensitivity and specificity of the SARS-CoV-2 IgG/IgM diagnostic classifier in NAT+ subjects and pre-pandemic controls. Z-scores for each motif were summed to generate an IgG/IgM composite score. The maximum value for the IgG or IgM for each sample is shown. Samples above a cutoff of 25 are classified as positive. The sensitivity or specificity of the SERA panels for all COVID-19 cohorts and controls is shown above each column. (**C**) Candidate structural mappings for the linear motif LPFQQ and non-mapping motif YWXYFXK (through its related variant YXXYFXK). Inset highlights the key contact amino acid residues for the proposed structural localizations of the motifs. (PDB codes: 7A95 and 6ZGG respectively). (**D**) Neutralization titer vs. log-enrichment for each motif (LFPQQ and YWXYFXK) shows that samples with the highest enrichment for either motif tend to have higher titer neutralization activity.

### SARS-CoV-2 diagnostic panels can classify NAT+ samples with sensitivity comparable to ELISA

To develop a SARS-CoV-2 diagnostic panel, we selected a subset of peptide motifs that, in the training cohort, either exhibited high sensitivity and specificity or improved the breadth of coverage (Supplemental Table 2). We normalized and summed motif enrichments to generate a composite score and compared sub-panels to identify the panel with the maximal diagnostic performance on the training cohort (Methods). A composite score of ≥25 was set as a cutoff for both IgG and IgM panels to obtain a specificity of ≥99% on the pre-pandemic training controls (Table 1). The panel performance was evaluated on a test cohort of 427 COVID-19 samples that were confirmed positive by NAT testing (Table 1, testing cohorts I-III). The classifier with the best overall performance is shown in Figure 3B. The sensitivity varied between 54% and 82% across the NAT+ cases from different cohorts, primarily based on the timing of blood collection relative to symptom onset for each cohort. A specificity of 99.3% for IgG and 99.1% for IgM was achieved on a test set of 1500 pre-pandemic repertoires that were tested for acute illness. Combining the IgG and IgM panels into a single test resulted in a panel specificity of 98.7%. Notably, no pre-pandemic samples were co-positive for IgG and M, thus the specificity for subjects that were positive for both IgG and IgM was 100% in the test control set. Forty-two percent of all tested COVID-19 samples met these criteria.

We plotted the SERA scores for samples from cohorts I and II, where timing of the blood draw relative to date of symptom onset was provided (Figure 3B). The panel exhibited a sensitivity of 47% at 0-5 days after symptom onset, 64% at 5-10 days and ≥90% at >10 days post symptom onset. Where predicate SARS-CoV-2 ELISA results were available, we compared performance relative to SERA in SARS-CoV-2 NAT+ samples. Overall, SERA IgG and IgM panels together demonstrated similar sensitivity to three different ELISAs in current use (95% CI, Wilson score) (Table 2).

**Table 2:**
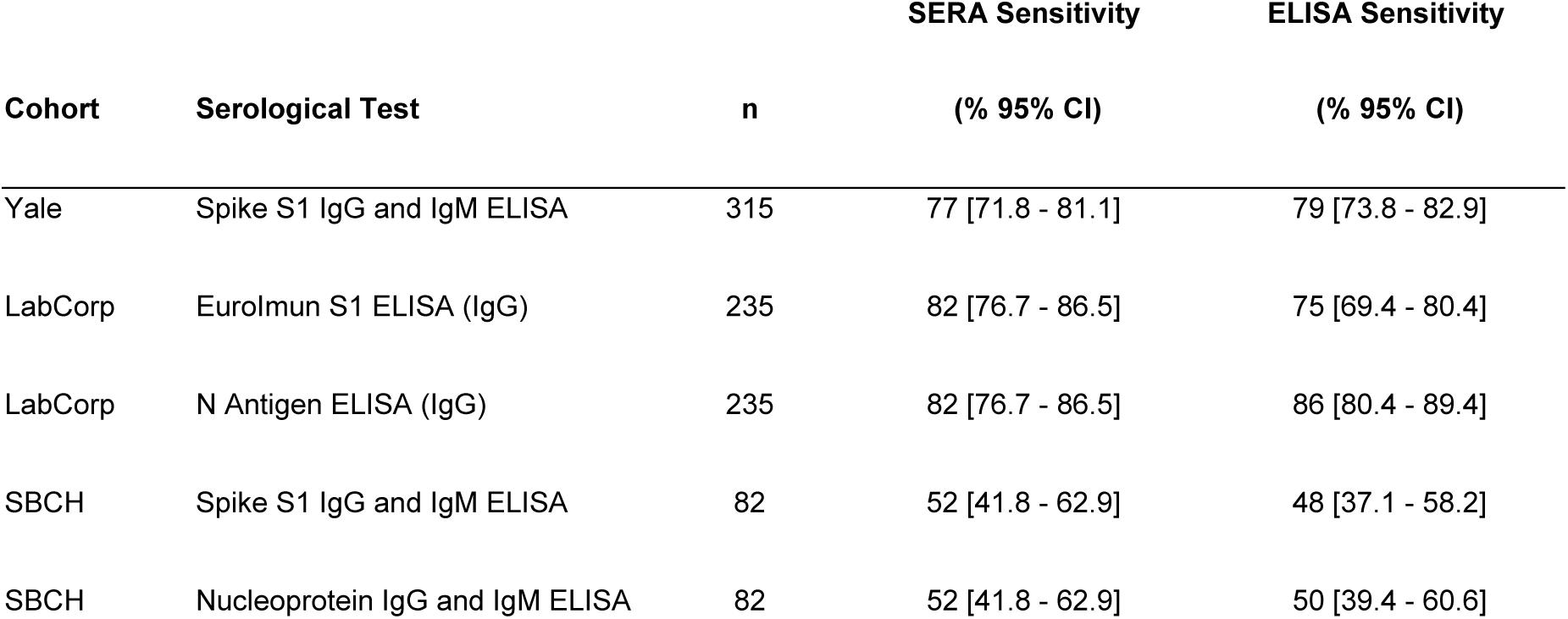
SERA SARS-CoV-2 IgG or IgM panel sensitivity as compared with various ELISAs in NAT positive subjects.

| Cohort | Serological Test | n | SERA Sensitivity | ELISA Sensitivity |
| --- | --- | --- | --- | --- |
|  |  |  | (% 95% CI) | (% 95% CI) |
| Yale | Spike S1 IgG and IgM ELISA | 315 | 77 [71.8 - 81.1] | 79 [73.8 - 82.9] |
| LabCorp | EuroImmun S1 ELISA (IgG) | 235 | 82 [76.7 - 86.5] | 75 [69.4 - 80.4] |
| LabCorp | N Antigen ELISA (IgG) | 235 | 82 [76.7 - 86.5] | 86 [80.4 - 89.4] |
| SBCH | Spike S1 IgG and IgM ELISA | 82 | 52 [41.8 - 62.9] | 48 [37.1 - 58.2] |
| SBCH | Nucleoprotein IgG and IgM ELISA | 82 | 52 [41.8 - 62.9] | 50 [39.4 - 60.6] |

### Structural epitope mapping

To further interrogate functional relevance of the identified motifs, we mapped motifs to the surface of published SARS-CoV-2 spike glycoprotein crystal structures. We observed that the linear motif LPFQQ, which has been previously implicated as a neutralizing epitope^25^, mapped to a single location on the surface of spike glycoprotein (Figure 3C). Using these same methods, we examined possible structural maps of motifs without linear maps to SARS-CoV-2. We highlight one exemplary motif, YWXYFXK which was found to map to the RBD of spike glycoprotein (Figure S2). Based on our previous observations that tryptophan tends to confer structural characteristics on our peptide epitopes, we additionally examined the mapping of the slightly modified motif YXXYFXK which we found also maps strongly to spike (Figure 3C). In addition to the highlighted match to spike, YXXYFXK had two less feasible maps to spike glycoprotein (Figure S2).

We also investigated the potential neutralization capacity of these motifs. We plotted the neutralization titer of each sample against the enrichment of the motif in those samples (Figure 3D). For both motifs, we observed that higher enrichment values tended to be present in patients with higher titer neutralization activity.

### Motifs and epitopes associated with disease severity

Based on prior studies that described subjects with severe disease possessing a stronger and, perhaps, earlier humoral IgG response in both spike and nucleoprotein relative to subjects with mild disease^29,30^, we examined differences in epitope severity detected by SERA. We compared the SERA IgG panel score (developed to distinguish COVID-19 patients from pre-pandemic controls) across the spectrum of severities present in our population (Figure 4A). We observed a significant elevation of the panel score in patients with severe or moderate disease compared to their mild disease counterparts. To understand the specific epitopes driving the severity delineation, we identified the 10 motifs with the most significant t-test p-value when comparing severe and mild disease (Figure 4B). We observe a potential confounding of days since onset of symptoms with the SERA IgG score (Figure S3). All 10 motifs were identified in the IgG screen and 9 out of 10 motifs did not possess a linear map to SARS-CoV-2. In the hierarchical clustering of samples, we observe subsets of severe patients with preferential enrichment for differing motifs. After splitting our data into 2/3 training and 1/3 testing cohorts, we built a simple LASSO model to classify moderate/severe from mild disease, and observed encouraging performance (training AUC 0.92, testing AUC 0.9, Figure S3).

**Figure 4:**
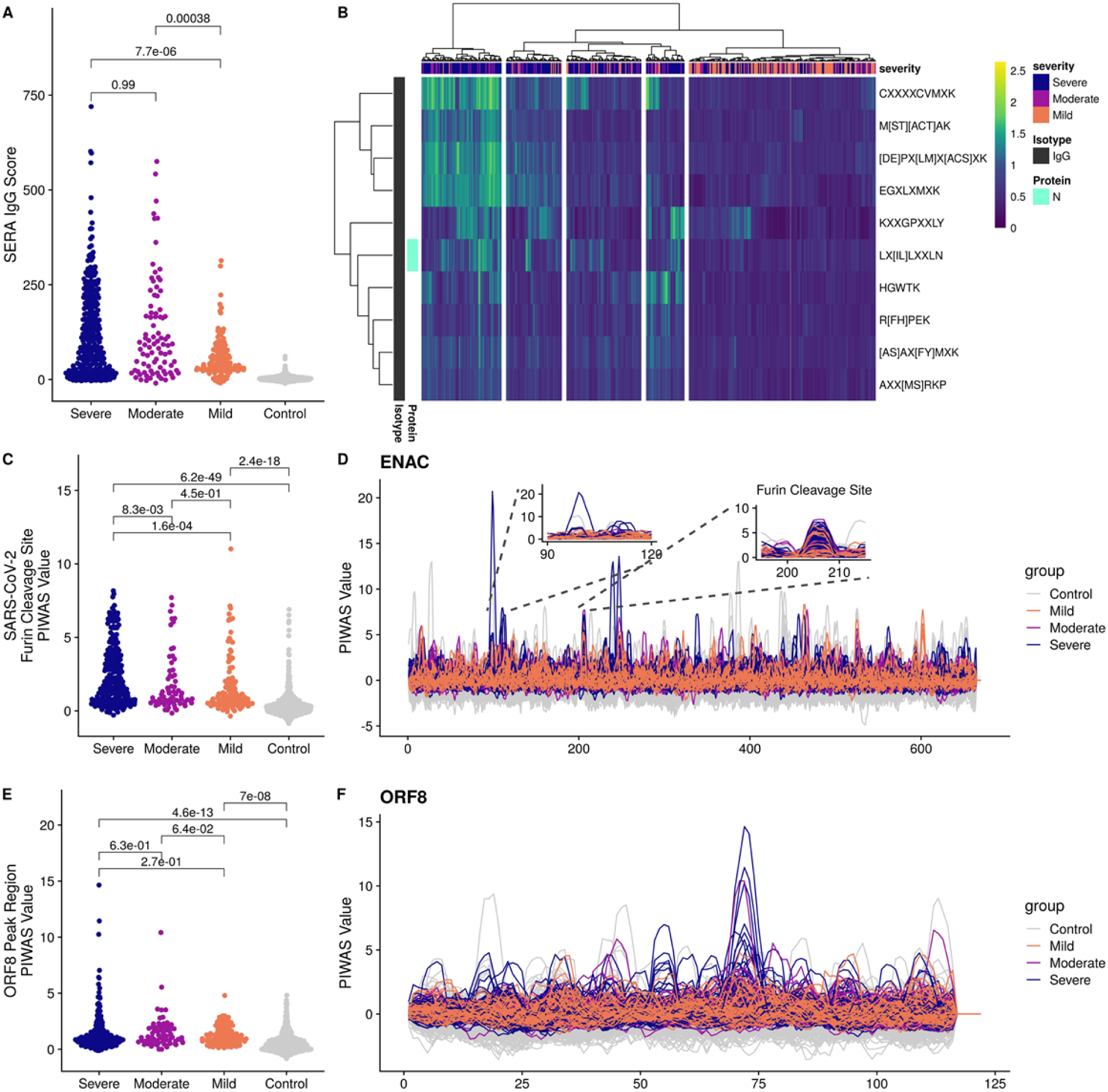
Significantly different epitope signals are observed in mild, moderate, and severe cases of COVID-19. (**A**) Comparison of SERA total IgG motif panel scores for severe, moderate, and mild cases based on motifs used in the diagnostic panel (Figure 3). Colors indicate disease severity. (**B**) Severe, moderate, and mild cases of SARS-CoV-2 are clustered based on log-enrichment of the top 10 motifs identified by a t-test comparison of severe and mild patients. (**C**) Distribution of PIWAS values at SARS-CoV-2 furin cleavage site for severe, moderate, and mild cases. (**D**) PIWAS tiling data is shown for the human ENaC-α protein. Insets highlight the furin cleavage site with homology to spike (right) and a non-homologous region of ENaC-α (left). (**E**) Distribution of PIWAS values for the peak epitope in ORF8 for severe, moderate, and mild cases. (**F**) PIWAS tiling of individual samples on the entire ORF8 sequence. All p-values were calculated using outlier sum statistical test.

One of the distinguishing features of the SARS-CoV-2 coronavirus is the acquisition of polybasic residues (RRAR) at the cleavage site of the S1/S2 boundary. Cleavage of spike protein at this site is required to enable viral membrane fusion^31,32^. It has been proposed that this novel sequence enables the virus to take advantage of host proteases, such as furin, that cleave proteins with this recognition sequence, thereby increasing the potential tropism of the virus relative to other coronaviruses^31,33^. We asked if this site elicited an immune response, and if so, was it seen differentially in subjects with different disease severity. In the spike epitope map, signal at this sequence location is both prominent and prevalent in the cohorts — 120 out of 385, or 31% of subjects, had epitope signals >99% of that seen in controls. We also determined that the site elicited a statistically significantly stronger immune response in subjects with severe disease relative to subjects with mild or moderate disease (Figure 4C). Specifically, 39%, 23%, and 20% of severe, moderate, and mild cases, respectively, had strong epitope signals greater than 99% of that in the controls.

The novel eight amino acid furin cleavage site (RRAR|SVAS) in spike maps identically to a peptide sequence in one protein in the human proteome, the amiloride sensitive sodium channel ENaC-α protein^33^. This protein is expressed on the surface of multiple tissues implicated in COVID-19 pathology, and similar to spike, requires cleavage for activation. As the sites share the eight amino acid furin cleavage sequence, not surprisingly, we see a highly correlated PIWAS immune signal in both proteins (Figure S3) that is also statistically stronger in severe disease relative to mild or moderate disease (Figure 4D). We also note that in severe cases, a number of very strong epitopes in ENaC-α outside of the cleavage site are seen relative to mild cases. The signal at both sites was also seen to increase over time, particularly between 2 and 4 weeks, indicating a likely adaptive immune response to this site (Figure S3).

In addition to spike and nucleoprotein, a robust immune response has been described against the ORF8 protein^34^. Several reports have described a variant of SARS-CoV-2 with a 382-nucleotide deletion in ORF7b and ORF8 as well as an association of the deletion with a milder disease course^35^. While we do not have genotype information for all strains, based on the GISAID database we assume that most of the samples in our cohorts do not have this deletion. To explore the possible association of immune response with disease severity, we analyzed the PIWAS signal against ORF8, which encompasses most of the 382-nucleotide deletion. While there appear to be more extremely high signals in severe cases, using an outlier sum statistic, the PIWAS signal in ORF8 does not reach statistical significance in severe cases relative to mild and moderate cases (Figure 4E, F).

### PIWAS prediction of antibody cross-reactivity to other coronaviruses

We next investigated SARS-CoV-2 epitopes that may cross-react with other coronaviruses as previous exposure to coronaviruses could have protective or even deleterious effects on symptoms^6,7^. To identify potential cross-reactive epitopes, we performed PIWAS using the epitope repertoires from COVID-19 samples against various coronavirus proteomes, including SARS-CoV-2, SARS-CoV (SARS), MERS, and the four common human coronaviruses (hCoVs) HKU1, OC43, 229E, and NL63. Analysis of average PIWAS values for spike glycoprotein across coronaviruses revealed epitopes that were conserved in many coronaviruses as well as epitopes that were specific to SARS-CoV-2 (Figure 5A). We identified 10 epitopes enriched against the SARS-CoV-2 proteome (average PIWAS >0.5), two and one of which overlapped with OC43 and NL63 epitopes, respectively. For example, the region corresponding to spike 809-834 in SARS-CoV-2 (alignment indices 1140-1170) contained an epitope that was observed against all coronaviruses analyzed (Figure 5B). However, at spike 1141-1162 in SARS-CoV-2 (alignment indices 1500-1525) an epitope was observed only against SARS-CoV-2, SARS, MERS, and OC43 proteomes, with OC43 exhibiting the highest average PIWAS value. After evaluating enrichment for these cross-reactive spike epitopes in COVID-19 cases with different disease severity, we found there was no statistical difference between severe, moderate, and mild cases (Figure 5C).

**Figure 5:**
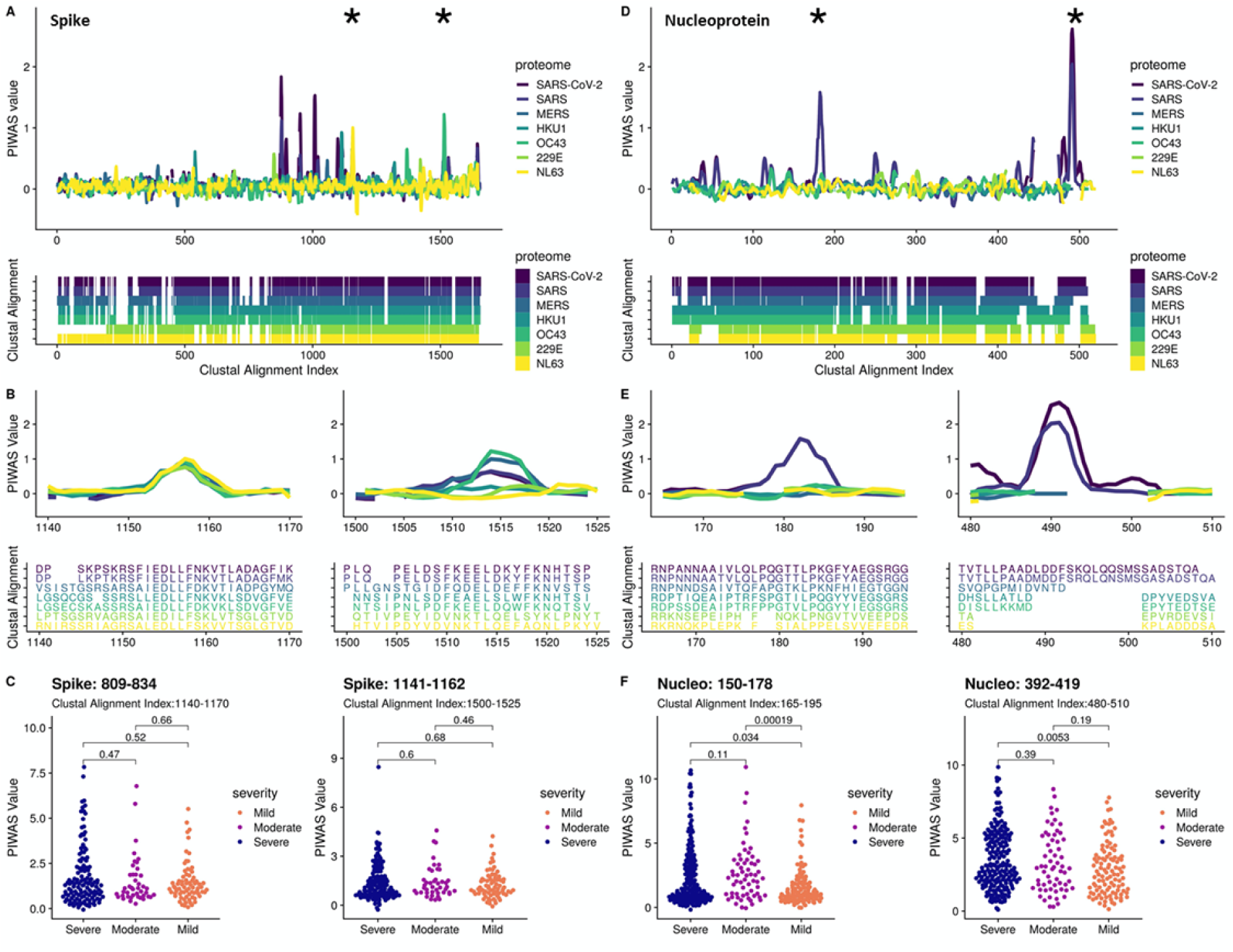
Cross-reactivity analysis across coronaviruses reveals shared epitopes and epitopes specific to SARS-CoV-2. PIWAS was performed using COVID-19 samples against various coronavirus proteomes including SARS-CoV-2, SARS, MERS, and the common hCoVs HKU1, OC43, 229E, and NL63. PIWAS tilings for (**A**) spike and (**D**) nucleoprotein revealed regions of cross-reactivity as well as epitopes only observed against SARS-CoV-2. Clustal multiple sequence alignments were performed and visualized below to depict sequence similarity and divergence. Distinct epitopes from (**B**) spike and (**E**) nucleoprotein showcase PIWAS values across the coronaviruses with corresponding clustal alignment sequences below. Epitope locations are denoted with asterisks in (**A**) and (**D**). Distribution of PIWAS values at epitopes from spike (**C**) and nucleoprotein (**F**) for severe, moderate, and mild cases, with p-values from Wilcoxon-rank sum test.

In contrast to spike, nucleoprotein exclusively contained epitopes specific to SARS-CoV-2 and SARS, with 4 epitopes against the SARS-CoV-2 proteome (Figure 5D). Strong epitopes were observed against SARS-CoV-2 at regions 150-178 (alignment indices 165-195) and 392-419 (alignment indices 480-510) with no signal observed against hCoVs (Figure 5E). We determined that these nucleoprotein epitopes were significantly enriched in severe and/or moderate cases compared to mild cases (Figure 5F).

### Epitope signal in mutated SARS-CoV-2 strains

To study the possible effects of known mutations to the SARS-CoV-2 virus on antibody response, we leveraged the ability of PIWAS to interrogate the SERA database with any sequence of interest. In the 96,437 sequenced strains from GISAID, we enumerated 21,127 distinct amino acid mutations to spike glycoprotein, nucleoprotein, envelope protein, and membrane protein^36,37^. For each mutation, we compared epitope signal against the wild-type (WT) and mutant position in every COVID-19 specimen. We observed a bias towards mutations yielding a decreased PIWAS signal relative to WT (Figure 6A). A subset of these mutations yielded decreased signal across a large number of COVID-19 patients (Figure 6B). To assess the significance of this decreased epitope signal, we *in silico* randomly mutated amino acids throughout the same protein sequences as a null distribution. The Kolmogorov-Smirnov test comparing the observed and null distributions was highly significant (p=3e-11), indicating that the bias towards mutants that generate a decreased epitope signal exceeds that which would be explained purely due to chance (Figure S4). For membrane protein, nucleoprotein, and spike glycoprotein, we highlight exemplar mutations which resulted in decreased epitope signal across a large number of patients (Figure 6C) and, in the case of spike glycoprotein, are on the surface of the protein according to the crystal structures considered in this paper^31,32^. In contrast, the dominant spike glycoprotein D614G exhibits no epitope signal for either the wild-type or mutant strains (Figure 6D).

**Figure 6:**
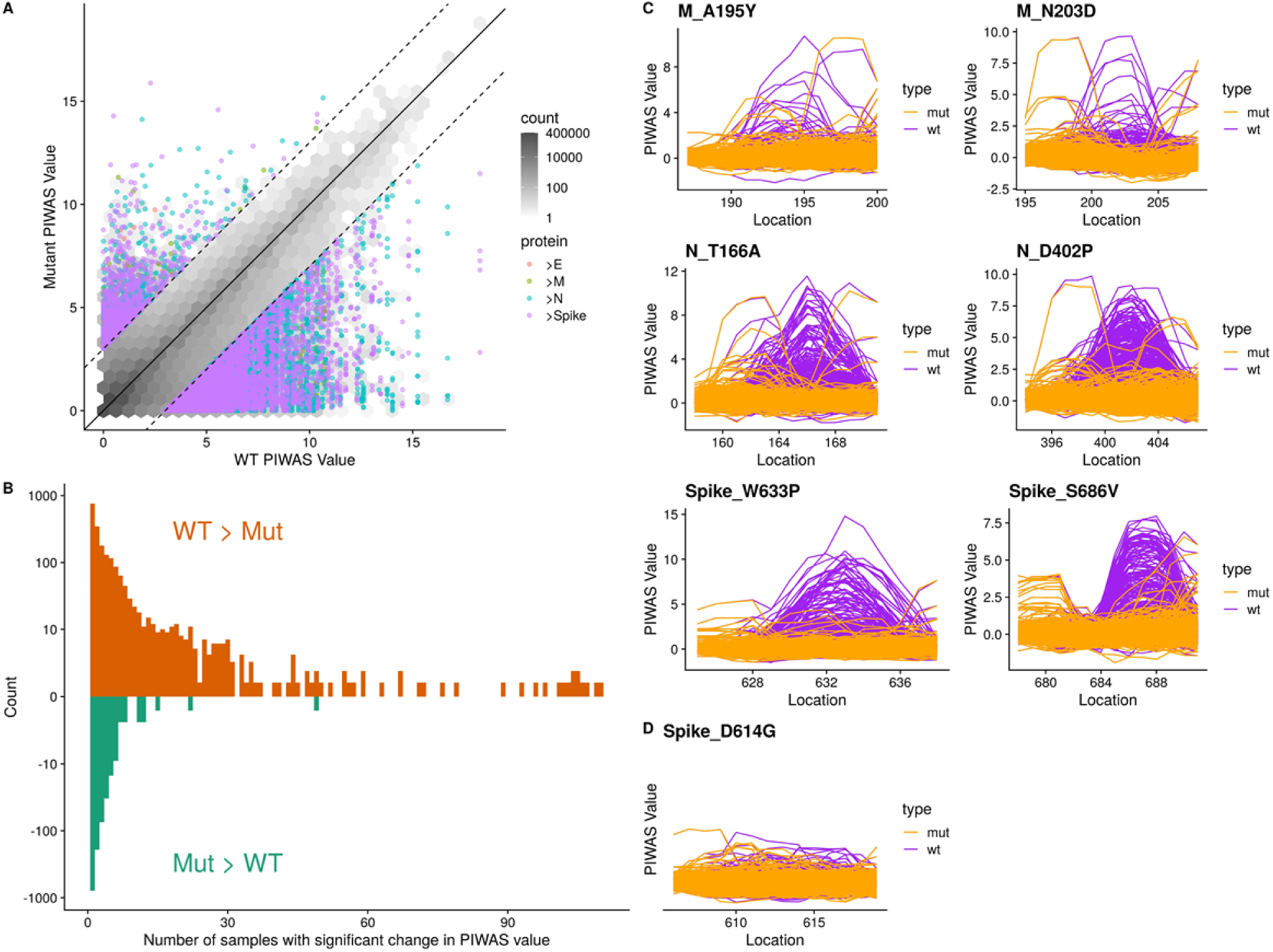
Mutations to SARS-CoV-2 are biased towards decreasing immune epitope response. 21,127 distinct amino acid mutations in spike glycoprotein, nucleoprotein, envelope protein, and membrane protein in SARS-CoV-2 strains were identified from sequencing data of 96,437 genomes from GISAID. (**A**) For each mutation, the PIWAS value of the wild-type (WT) sequence was compared to the PIWAS value for the mutated strain (mut). (**B**) Mutations conferring a significant PIWAS value change (|PIWAS_WT_-PIWAS_mut_| > 3) for each COVID-19 sample were identified. For each mutation, the number of patients with a significant difference was counted. (**C**) Exemplary mutations that yielded a decrease in PIWAS values are shown for membrane protein (top row), nucleoprotein (middle row), and spike (bottom row). (**D**) No significant immune signal is seen at location 614 of spike, for either the wild-type or the D614G variant.

## Discussion

While conventional serology is a cornerstone of infectious disease diagnosis, the COVID-19 pandemic has raised many questions not answered by these testing modalities alone. Here we have shown that high-content random bacterial peptide display library screening using SERA provides a tool to broadly and deeply probe individual antibody repertoires. These profiles, both individually and in the aggregate, can yield insights into disease severity, immunity, cross-reactivity to other coronaviruses (including SARS-CoV-2 mutant strains), and autoimmune sequelae.

By taking a focused, proteome-constrained approach to identifying signal against the SARS-CoV-2 proteome, we both reiterate the established immunological relevance of spike and nucleoprotein as well as identify less described signals against protein 3a and NSP-8. Epitope-level characterization of these antigens highlights particularly immunogenic epitopes within each protein, which might serve as targets in the development of vaccines and therapeutics. In particular, we identified strong epitopes in nucleoprotein including at amino acids 158-172 and the C-terminal domain 380-419, as well as epitopes in spike glycoprotein including at amino acids 555-572, 810-828, and 1145-1159, consistent with previous studies^4,24,38^. Additionally, we highlight novel and less-studied epitopes, including a dominant IgM epitope from membrane protein (amino acids 1-12) which could provide utility in early diagnostics. While we did not observe spike RBD epitopes that were conserved across the patient cohort, we found compelling examples of private RBD epitopes. The lack of linear epitopes towards spike RBD is unsurprising given the complex structural nature of spike, with many strands running in parallel likely yielding an abundance of structural mimotopes (also reflected in the quantity of non-mapping motifs in our diagnostic panel).

Leveraging our database of thousands of pre-pandemic repertoires collected from healthy individuals as well as people with infections, autoimmune diseases, and cancer across all age groups and geographies, we were able to assess the specificity of the SARS-CoV-2 antibody response and identify a panel of epitopes that could distinguish COVID-19 cases from controls with accuracy similar to conventional serological testing.

We further investigated the possible origins of the non-mapping motifs by attempting to map them structurally to spike glycoprotein. We validated the mapping by showing that it accurately identifies the linear motif LPFQQ, and then applied the method to non-mapping motifs. We find that the motif YWXYFXK exclusively maps to the RBD. However, previous observations have suggested that tryptophan (W) may be more important for conferring structure to the mimotope than for identity mapping, yet still the more general motif YXXYFXK maps to RBD as well. When combined with the neutralization titers of samples in which this motif is enriched, it is possible to speculate about the mechanism of neutralization. If the antibodies that recognize this motif bind to the RBD of spike glycoprotein, they may block the ability of SARS-CoV-2 to bind to ACE2 and inhibit viral entry and infection.

One of the ongoing areas of development in this approach is that while we have assumed that the identity of the residues remains constant between motif and the epitope, it is possible that amino acid substitutions could be allowed. We have attempted a first pass to mitigate this by allowing for a more general mapping with the modification of the W to an X, but we continue to iterate on this model to more accurately account for residue mismatches in mapping motifs to the structure. While these methods are still under development, the results here demonstrate the applications of such a method.

Consistent with previous studies, we find that the humoral immune response against SARS-CoV-2 is stronger in severe and moderate disease relative to mild disease^4,39,40^. This finding is consistent with a general pattern of disease associated with immunopathology in COVID-19. We also identified specific epitope profiles that correlate with disease severity and combined these epitopes into a preliminary disease severity classification model. To further validate these findings we would require a separate, validation cohort of patients which span mild and severe disease states. Importantly, many of our disease severity analyses are potentially confounded by the number of days since onset of symptoms in the COVID-19 subpopulations, partially due to the challenge of both identifying disease onset in mild patients and collecting samples from non-hospitalized patients.

Using *in silico* analysis of repertoires on the human proteome, we are also able to identify candidate cross-reactive or novel autoantigen epitopes that may be important in disease pathogenesis. The polybasic cleavage site seen in SARS-CoV-2 is unique among coronaviruses and potentially enables it to increase its tissue tropism^33^. We demonstrate that the immune response at this site is predicted to be both significantly prominent and prevalent relative to a pre-pandemic cohort, as well as significantly stronger in severe and moderate versus mild disease. The sequence is shared with the amiloride sensitive sodium channel ENaC-α, which is responsible for sodium influx into tissue^41^. Disruption of this channel in individuals with variants has been associated with pulmonary edema and alveoloar fluid overload^40,42^. Thus, this channel has been investigated in SARS-CoV-2 and other respiratory illnesses for possible pathogenic implication in disease and as a target for therapy^43^.

Our finding that immune signal at this site is significantly elevated suggests the possibility of molecular mimicry with potential pathogenic consequences. While we do not have direct evidence of binding of antibody to ENaC-α at this site, functional blockage of the site could be postulated to prevent the cleavage of the ENaC-α channel, which is necessary for channel activation^43^. The signal against this site increases over time in the majority of cases indicating an adaptive immune response, but the effect of this response on pathogenicity is unclear as binding at the spike site could in fact reduce infectivity of the virus. A recent study has also noted that binding at this site reduces the RBD-ACE2 binding energy, and therefore could be a potential site for neutralizing antibodies^44^. Functional analyses and experiments are thus required to investigate whether cross-reactivity occurs and to distinguish the effect of antibodies binding to either the viral or host antigens at this site. The ability to query potential autoantigen signal using SERA in the context of SARS-CoV-2 infection and epitopes is an area for continued inquiry given the mounting data supporting the significance of autoantigens in the immunopathology of COVID-19^10,45^.

Milder disease has been described in subjects with the 382-ORF8 deletion variant, and the ORF8 protein has been noted to be associated with strong humoral response^35,46^. In our study, we also see significant response relative to a pre-pandemic cohort in ORF8. While a few epitopes appear quite strong in some individuals in ORF8 with severe disease, the overall signal across the antigen was not seen to be statistically significant in mild versus severe disease. Specific, strong epitope signals in ORF8 could be postulated to contribute to severe disease through a variety of mechanisms, but this would also need to be explored through further epidemiological and experimental analysis.

By evaluating epitope signal in COVID-19 cases against common human coronavirus (hCoV) proteomes, we predicted prevalent cross-reactive epitopes particularly in the S2 domain of spike. Given the strength and prevalence of these cross-reactive epitopes, it is plausible that previous exposure to hCoVs contributed to these antibody responses, a boosting phenomenon recently described in COVID-19 cases^47^. In particular, the cross-reactive epitope at spike amino acids 809-834 near the fusion domain has been shown to elicit an antibody response in SARS-CoV-2 uninfected adolescents and adults^48^. Interestingly, antibodies targeting this epitope demonstrated neutralizing capacity using antibody depletion assays^38^. More broadly, the presence of spike-reactive T cells in healthy donors has been observed against SARS-CoV-2 as well as hCoVs 229E and OC43, primarily reactive towards the spike S2 domain^6^. While these findings suggest a role for cross-reactive epitopes in the response to SARS-CoV-2 infection, it is uncertain what impact pre-existing antibodies have towards protection, immunity, and disease progression. Recent studies suggest that pre-existing antibodies from hCoVs exist but are not associated with protection^47,49^. We observed that prevalent cross-reactive epitopes in spike were not associated with COVID-19 severity while multiple nucleoprotein epitopes specific to SARS-CoV-2 and SARS were significantly enriched in severe cases compared to mild. Notably, it has been shown that convalescent COVID-19 patients exhibited a shift in antibody response towards spike compared to a nucleoprotein-directed antibody response in deceased patients^3^. Given that cross-reactive epitopes were observed in spike, additional investigation will be critical towards understanding pre-existing antibody responses that may impact SARS-CoV-2 infection and COVID-19 progression.

The decreased epitope signal in COVID-19 patients against mutant strains of SARS-CoV-2 compared to WT is suggestive of evolutionary evasion of the antibody response^50–52^. Under this hypothesis, SARS-CoV-2 strains are undergoing selective pressure to evade antibody response, resulting in strains that may be less susceptible to clearance by those previously infected with wild-type SARS-CoV-2^53,54^. While these observations do not inherently indicate greater infectivity, the public health consequences of epitope mutation are concerning and further suggestive of the potential for the SARS-CoV-2 virus to periodically re-emerge and reinfect individuals with prior exposure. While not conclusive, these observations reveal the importance of monitoring epitope mutations and could be used to guide therapeutic and vaccine development efforts to focus on epitopes that are less susceptible to evasion, which would be more broadly cross-reactive and robust to evolutionary changes.

The dominant strain of SARS-CoV-2 which is now in circulation possesses the D614G mutation. Based on our data, neither the wild-type nor the mutant confer a significant linear epitope, consistent with observations that the mutation is most notable for its effect on the structure of spike^55^.

We acknowledge various limitations with the SERA platform that impact this study. Much of this study has focused on dominant epitopes prevalent in COVID-19 cases, but many of the private epitopes not explicitly discussed here, particularly in spike RBD, are critical to fully understanding the protective antibody response and clinical outcomes. Moreover, there are clear limitations for probing the epitope repertoire with linear peptides, chiefly the challenges of identifying structural epitopes and the role of post-translational modifications such as glycans^56^. While a random peptide library enables unique opportunities to identify structural mimics, much work remains in cataloguing and mapping these mimics to their cognate antigens.

In summary, we present the application of SERA to assess SARS-CoV-2 seropositivity and to characterize a high-resolution map of motifs and epitopes in individuals and populations. We demonstrate the ability of the platform to assess disease severity, to identify structural motifs associated with neutralization, to compare *in silico* epitope response to multiple coronavirus strains, to assess potential immune escape at sites of variation, to evaluate longitudinal changes in signal, and to reveal potential autoantigen response, all with one assay. The random nature of the libraries, the ability to identify linear mimics of structural epitopes, and the ability to leverage quality-controlled reference data from a large pre-pandemic cohort all contribute to SERA’s ability to elucidate the humoral immune response in SARS-CoV-2 infection.

Our findings support those of other studies that find clear differences in the humoral response of individuals with different clinical severity and trajectories. While we may identify associations between high resolution epitope and motif signals and disease severity, much work is required to establish functional or causal relationships. Examining and correlating epitopes to clinical efficacy in the context of vaccines and therapeutic antibodies will help to elucidate the connection between measured immune response and patient outcome.

Yet the epitope landscape can change, as it is already clear that coronaviruses mutate and SARS-CoV-2 is no exception. Potential changes in the infectivity of the virus in just this first year of the current health crisis underscore the need to track evolving immune responses and clinical features in populations world-wide^55,57–60^. We have demonstrated the ability to capture and query both past and present repertoires through analysis of both pre-pandemic and current pandemic samples. Using SERA to observe longitudinal immune responses against the human proteome and coronavirus in the context of persistent symptoms or reinfection enables construction of a detailed picture of infection, immunity and disease in COVID-19. SERA’s ability to query against any variant or future emerging genomes can be used to support ongoing management of the current health crisis and future novel outbreaks.

## Materials and Methods

### Biospecimens and Cohorts

Sera or plasma from confirmed or suspected COVID-19 cases were acquired from Yale, Santa Barbara Cottage Hospital (SBCH), LabCorp, BioIVT and Blood Centers of America (BCA). Samples were de-identified prior to receipt at Serimmune. All samples and associated metadata are shown in Supplemental Table 1.

### Yale cohort

Samples were acquired as part of the Yale IMPACT biorepository study. The cohort included inpatients that tested positive by PCR for SARS-CoV-2, outpatients with suspected disease based on symptoms and Health Care Workers (HCW) that became positive by serial PCR or serology testing every 2 weeks. COVID-19 cases were classified as mild if patients were not hospitalized, moderate if hospitalized, and severe if on high-flow nasal canula, BiPAP or other non-invasive ventilation, intubated or died from COVID-19. Participation in the study was voluntary and the study protocol was approved by the Yale Institutional Review Board.

### SBCH cohort

Biobanked sera or plasma from individuals that previously tested positive for COVID-19 were provided by the SBCH Biobank. Clinical data, including age, sex, and disease severity were obtained by SBCH staff for inclusion in the biobank. Specimens were collected from both inpatient and ambulatory settings and were coded as asymptomatic, mild/moderate if the subject had symptoms consistent with COVID-19, or severe if the individual required admission to the ICU for symptoms. Participation in these studies was voluntary and the study protocol was approved by the SBCH Institutional Review Board.

### LabCorp cohort

The majority of samples were remnant sera from acutely ill, ICU hospitalized, PCR confirmed COVID-19 cases with high IL-6 test results (n=235). These cases were classified as severe disease. An additional ten suspected COVID-19 cases were from individuals with mild symptoms. A subset of these had serological evidence of infection by anti-RBD ELISA and/or neutralization assay data.

### BioIVT cohort

Remnant serum samples with serological evidence of infection by a positive Epitope EDI IgG test (n=20) or a positive NAT test (n=1) were purchased from BioIVT. A subset had disease severity characterization provided by the vendor.

### BCA cohort

Plasma samples were collected from healthy blood donors in New York during the period of March–July of 2020 as part of a collaboration with The Blood Centers of America. Two samples included in the study were collected from COVID-19 plasma donors with confirmed disease. Suspected COVID-19 cases included in the study had serological evidence of infection based on a positive SERA IgG or IgM result that was subsequently confirmed by S1 spike and nucleocapsid ELISA IgG in the majority of cases. Cases from healthy donors were classified as mild disease.

### SERA serum screening

A detailed description of the SERA assay has been published^26^. For this study, serum or plasma was incubated with a fully random 12-mer bacterial display peptide library (1×10^10^ diversity, 10-fold oversampled) at a 1:25 dilution in a 96-well, deep well plate format. Antibody-bound bacterial clones were selected with 50 µL Protein A/G Sera-Mag SpeedBeads (GE Life Sciences, cat#17152104010350) (IgG) or by incubation with a biotinylated anti-human IgM antibody (Jackson ImmunoResearch, cat# 709-066-073) final assay dilution 1:100, followed by a second incubation with 50 ul Dynabead MyOne Streptavidin T1 conjugated magnetic beads (IgM) (Thermo-Fisher 65602). The selected bacterial pools were resuspended in growth media and incubated at 37°C shaking overnight at 300 RPM to propagate the bacteria. Plasmid purification, PCR amplification of peptide encoding DNA, barcoding with well-specific indices was performed as described ^26^. Samples were normalized to a final concentration of 4nM for each pool and run on the Illumina NextSeq500.

### Spike S1 and nucleoprotein ELISA

The SARS-CoV-2 Spike S1 and N antigen ELISA data were provided by Yale and LabCorp. Spike S1 and nucleoprotein ELISAs on the SBCH COVID-19 samples were performed in house using recombinant proteins (ACRO Biosystems, S1N-C52H3 and NUN-C5227, respectively). A cut-off value for positivity was established using 3 times the standard deviation of 502 pre-pandemic controls for the IgG and 82 pre-pandemic controls for IgM assays. Briefly, plates (Nunc MaxiSorp) were coated with recombinant proteins, 0.5 ug/ml for IgG and 1 ug/ml for IgM at 4°C overnight. After washing, plates were blocked with PBS containing 5% non-fat milk for 2 hours at room temperature. Plates were then incubated with serum diluted 1/250 in blocking buffer for 1 hour at room temperature. Plates were washed, then incubated with HRP-goat anti-human IgG or HRP-donkey anti-human IgM (Jackson ImmunoResearch) secondary antibody diluted 1/10,000 in blocking buffer for 1 hour at room temperature. After washing, the reaction was developed with 3,3’,5,5’-teramethylbenzidine substrate solution (ThermoFisher) for 15 minutes and stopped with 1M HCL. The absorbance was measured on a Tecan Spectrafluor plus plate reader at 450 nm.

### Cell lines and virus

VeroE6 kidney epithelial cells) were cultured in Dulbecco’s Modified Eagle Medium (DMEM) supplemented with 1% sodium pyruvate (NEAA) and 5% fetal bovine serum (FBS) at 37°C and 5% CO2. The cell line was obtained from the ATCC and has been tested negative for contamination with mycoplasma. SARS-CoV-2, strain USA-WA1/2020, was obtained from BEI Resources (#NR-52281) and was amplified in VeroE6 cells. Cells were infected at a MOI 0.01 for four three days to generate a working stock and after incubation the supernatant was clarified by centrifugation (450g × 5min) and filtered through a 0.45-micron filter. The pelleted virus was then resuspended in PBS then aliquoted for storage at − 80°C. Viral titers were measured by standard plaque assay using Vero E6 cells. Briefly, 300ul of serial fold virus dilutions were used to infect Vero E6 cells in MEM supplemented NaHCO3, 4% FBS 0.6% Avicel RC-581. Plaques were resolved at 48hrs post infection by fixing in 10% formaldehyde for 1 hour followed by with 0.5% crystal violet in 20% ethanol staining. Plates were rinsed in water to plaques enumeration. All experiments were performed in a biosafety level 3 with the Yale Environmental Health and Safety office approval.

### Neutralization assay

Patient and healthy donor sera were isolated as before and then heat treated for 30m at 56 °C. Sixfold serially diluted plasma, from 1:3 to 1:2430 were incubated with SARS-CoV-2 for 1 h at 37 °C. The mixture was subsequently incubated with VeroE6 cells in a 6-well plate for 1hour, for adsorption. Then, cells were overlayed with MEM supplemented NaHCO_3_, 4% FBS 0.6% Avicel mixture. Plaques were resolved at 40hrs post infection by fixing in 10% formaldehyde for 1 hour followed by staining in 0.5% crystal violet. All experiments were performed in parallel with negative controls sera, at an established viral concentration to generate 60-120 plaques/well.

### PIWAS analysis

We applied the previously published PIWAS method^28^ to identify antigen and epitope signals against the Uniprot reference SARS-CoV-2 proteome (UP000464024)^61^. The PIWAS analysis was run on the IgG SERA data with a single sample per COVID-19 patient (for a total of 579 patients) versus 497 discovery pre-pandemic controls, and the 1500 validation controls used as the normalization cohort. Additional parameters include: a smoothing window size of 5 5mers and 5 6mers; z-score normalization of kmer enrichments; maximum peak value; and generation of epitope level tiling data. Antigens were ranked using the Mann-Whitney U false discovery rate, following the hypotheses of conserved epitopes in the context of infectious disease. For top antigens, tiling data was generated for every case and control sample. 95^th^ quantile bands were calculated based on each population separately. The most significant RBD epitopes were identified in COVID-19 patients with draws from at least 2 timepoints and a PIWAS value of at least 6 occurring between the 319^th^ and 541^st^ amino acids.

### IMUNE-based motif discovery

Peptide motifs representing epitopes or mimotopes of SARS CoV-2 specific antibodies were discovered using the IMUNE algorithm^27^. A total of 164 antibody repertoires from 98 hospitalized subjects from the Yale IMPACT study were used for motif discovery. The majority of subjects were confirmed SARS CoV-2 positive by nucleic acid testing (NAT). IMUNE compared ∼30 disease repertoires with ∼30 pre-pandemic controls and identified peptide patterns that were statistically enriched (p value ≤ 0.01) in ≥25% of disease and absent from 100% of controls. Multiple assessments were run with different subsets of cases and controls both for IgG and IgM. Peptide patterns identified by IMUNE were clustered using a PAM30 matrix and combined into motifs. The output of IMUNE included hundreds of candidate IgG and IgM motifs. A motif was classified as positive in a given sample if the enrichment was ≥4 times the standard deviation above the mean of the training control set. The candidate motifs were further refined based on at least 98% specificity. The final set of motifs was validated for sensitivity and specificity on an additional 1500 pre-pandemic controls and 406 unique confirmed COVID-19 cases from four separate cohorts (test cohorts I-IV, Table 1).

### Development of a diagnostic classifier for COVID-19

To generate a diagnostic score that classified subjects as serologically positive for antibodies to COVID-19, motif enrichment values were normalized using the mean and standard deviation of enrichments within the training set of pre-pandemic control repertoires. Individual SARS-CoV-2 motif normalized “z-scores” were then summed to obtain a composite score for each sample. A composite score of 25 was established as a cutoff for positivity for each panel to obtain a specificity of >99% on the pre-pandemic training controls.

### Structural motif mapping

Structural motif mapping was carried out by identifying a network of neighboring residues on the surface of a protein structure and looking in that network for matches to the motif of interest. Neighboring residues were residues which had α-carbons within 8 Å. The surface of the protein structure was calculated using the MSMS program^62^ with a probe radius of 2.5 Å. These values are line with other algorithms for mapping mimotopes to structures^63,64^ and were further optimized using our in-house dataset of monoclonal antibodies (data not shown).

Once each match to a motif was found in the surface network of neighbors, each residue is scored by the number of ways it was found to match to a motif. For example, the motif SE[RI] might map to SER on the surface, and additionally to SEI on the surface, where the same S and E are used. In this case, S would have a value of 2, E would have a value of 2, while R would get a value of 1, and I would get a value of 1. Each residue was then colored according to these values to produce the heat maps shown in the figures.

Structures used were downloaded from rcsb.org^65,66^. Due to the recent focus on SARS-CoV-2 there are many structures for spike glycoprotein in PDB. The following structures were chosen due to their high sequence coverage, and in order to represent a variety of conformations and binding states to ACE2: 6ZGG,6ZGI,7A93,7A95,7A97^31,32^. PDB structures were processed with Biopython^67,68^ and visualized with PyMol^69^.

### Mild versus severe disease analysis

For samples where clinical severity was known, we compared SERA IgG panel scores using the outlier sum statistic^28,70^. Using a t-test, we compared enrichments for all IgG and IgM motifs between the severe/moderate and mild populations. The 10 most significant motifs were highlighted and hierarchically clustered (Euclidean distance, Ward clustering^71^). Severity based on PIWAS signal against the furin cleavage and ORF8 regions was similarly compared using the outlier sum statistic. To identify potential auto-antigenic signal against ENAC-α, a PIWAS analysis was performed (using the same parameters as above) using the Uniprot^61^ reference human proteome (UP000005640).

### Common coronavirus analysis

We identified Uniprot reference proteomes for the four common human coronaviruses [OC43 (UP00007552), HKU1 (UP000122230), 229E (UP0006716), and NL63 (UP000145724)] and more severe strains [SARS (UP000000354) and MERS(UP000171868)]^61^. For each proteome, we ran a PIWAS with the same parameters as the SARS-CoV-2 PIWAS (above). For spike glycoprotein and nucleoprotein, we averaged PIWAS tiling values for the COVID-19 cohort across each proteome. A multiple sequence alignment of all these coronavirus sequences was performed using Clustal Omega^72^. Using this alignment index, we identified regions of divergent and convergent signal across the coronavirus proteomes in the COVID-19 population. For regions of interest, we calculated the significance of differences in patient severity using the Wilcoxon rank sum test.

### GISAID originating laboratories

Proteome sequences of 96,437 SARS-CoV-2 strains were downloaded from the GISAID database. We gratefully acknowledge the authors from the originating laboratories responsible for obtaining these specimens and the submitting laboratories where genetic sequence data were generated and shared via the GISAID initiative^36,37^. Supplemental Table 3 provides a complete list of these strains, authors, and laboratories used in this manuscript (Supplemental Table 3).

### SARS-CoV-2 strain analysis

For each of the 96,437 SARS-CoV-2 proteomes, we identified amino acid mutations relative to the original SARS-CoV-2 strain (hCoV-19/Wuhan/WIV04/2019). Incomplete proteomes were not considered. A total of 21,127 unique amino acid mutations were identified across spike glycoprotein, membrane protein, envelope protein, and nucleoprotein. For each mutation, a region of 10 flanking amino acids on either side was considered as the mutated region, for comparison against the same length wild-type region. For every sample, we calculated and compared PIWAS scores for the wild-type and mutant sequences. To assess significance of the observed bias, we generated *in silico* random mutations to these same proteins and performed the same analysis. We compared the actual and random signals using a Kolmogorov-Smirnov test.

## Data Availability

Motif enrichment and PIWAS data have been made available at:
https://www.serimmune.com/covidData.zip

https://www.serimmune.com/covidData.zip

## Data availability statement

Motif enrichment and PIWAS data have been made available at: https://www.serimmune.com/covidData.zip

## Acknowledgements

We would like to thank the entire team at Serimmune who has made this study feasible, including: Jack Coupart, Cameron Gable, Timothy Johnston, Steve Kujawa, Ulysses Leyva, Vikram Mahuvakar, Noah Nasser, Kenyon Plummer, Heidi Smith, Elizabeth Stewart.

## Author contributions

Conceptualization: WAH, KK, JB, AI, AIK, JCS; Data generation: WAH, KK, JB, EBJ, MC, ACM, CSDC, SFF, LF, JF, GJ, DK, JK, CL, LLL, MCM, LP, JR, RW, EAW, Yale IMPACT Team; Data curation: WAH, KK, JB, EBJ, ACM, LF, DK, JK, CL, LP, AAW; Formal analysis: WAH, KK, JB, AD, MJ, JK, BM, JR, MZ, JCS; Resources: WAH, KK, JB, EBJ, MC, ACM, PSD, CSDC, AD, SFF, LF, JF, MJ, GJ, DK, JK, CL, LLL, BM, MCM, LP, JR, JRS, RW, EAW, MZ, AI, AIK, JCS; Visualization: WAH, KK, JB, MJ, BM, MZ, JCS; Writing-original draft: WAH, KK, JB, MJ, BM, MZ, JCS; Writing-reviewing & editing: WAH, KK, JB, EBJ, MC, ACM, PSD, CSDC, AD, SFF, LF, JF, MJ, GJ, DK, JK, CL, LLL, BM, MCM, LP, JR, JRS, RW, EAW, MZ, AI, AIK, JCS

## References

1. Nishiura, H. et al. Estimation of the asymptomatic ratio of novel coronavirus infections (COVID-19). International Journal of Infectious Diseases 94, 154–155 (2020).

2. Mizumoto, K., Kagaya, K., Zarebski, A. & Chowell, G. Estimating the asymptomatic proportion of coronavirus disease 2019 (COVID-19) cases on board the Diamond Princess cruise ship, Yokohama, Japan, 2020. Eurosurveillance 25, (2020).

3. Atyeo, C. et al. Distinct Early Serological Signatures Track with SARS-CoV-2 Survival. Immunity 53, 524-532.e4 (2020).

4. Amrun, S. N. et al. Linear B-cell epitopes in the spike and nucleocapsid proteins as markers of SARS-CoV-2 exposure and disease severity. EBioMedicine 58, (2020).

5. Mathew, D. et al. Deep immune profiling of COVID-19 patients reveals distinct immunotypes with therapeutic implications. Science 369, (2020).

6. Braun, J. et al. SARS-CoV-2-reactive T cells in healthy donors and patients with COVID-19. Nature (2020). doi:10.1038/s41586-020-2598-9

7. Mateus, J. et al. Selective and cross-reactive SARS-CoV-2 T cell epitopes in unexposed humans. Science (80-.). eabd3871 (2020). doi:10.1126/science.abd3871

8. Ehrenfeld, M. et al. Covid-19 and autoimmunity. Autoimmunity Reviews 19, (2020).

9. Gruber, C. et al. Mapping Systemic Inflammation and Antibody Responses in Multisystem Inflammatory Syndrome in Children (MIS-C). Cell (2020). doi:10.1016/j.cell.2020.09.034

10. Bastard, P. et al. Auto-antibodies against type I IFNs in patients with life-threatening COVID-19. Science (80-). 370, (2020).

11. Kreye, J., Reincke, S. M. & Prüss, H. Do cross-reactive antibodies cause neuropathology in COVID-19? Nature Reviews Immunology (2020). doi:10.1038/s41577-020-00458-y

12. Guilmot, A. et al. Immune-mediated neurological syndromes in SARS-CoV-2-infected patients. J. Neurol. (2020). doi:10.1007/s00415-020-10108-x

13. Belongia, E. A. & Osterholm, M. T. COVID-19 and flu, a perfect storm. Science (2020). doi:10.1126/science.abd2220

14. Servick, K. Coronavirus creates a flu season guessing game. Science 369, 890–891 (2020).

15. Marshall, M. The lasting misery of coronavirus long-haulers. Nature 585, (2020).

16. Nath, A. Long-Haul COVID. Neurology 95, 559–560 (2020).

17. Perego, E. et al. Why the Patient-Made Term ‘Long Covid’ is needed. Wellcome Open Res. 5, (2020).

18. Ibarrondo, F. J. et al. Rapid Decay of Anti–SARS-CoV-2 Antibodies in Persons with Mild Covid-19. N. Engl. J. Med. (2020). doi:10.1056/nejmc2025179

19. Edridge, A. W. D. et al. Seasonal coronavirus protective immunity is short-lasting. Nat. Med. (2020). doi:10.1038/s41591-020-1083-1

20. Shaman, J. & Galanti, M. Will SARS-CoV-2 become endemic? Science (80-.). (2020). doi:10.1126/science.abe5960

21. Kissler, S. M., Tedijanto, C., Goldstein, E., Grad, Y. H. & Lipsitch, M. Projecting the transmission dynamics of SARS-CoV-2 through the postpandemic period. Science (80-.). 368, 860–868 (2020).

22. Long, Q. X. et al. Antibody responses to SARS-CoV-2 in patients with COVID-19. Nat. Med. (2020). doi:10.1038/s41591-020-0897-1

23. Wajnberg, A. et al. Robust neutralizing antibodies to SARS-CoV-2 infection persist for months. Science (80-.). (2020). doi:10.1126/science.abd7728

24. Shrock, E. et al. Viral epitope profiling of COVID-19 patients reveals cross-reactivity and correlates of severity. Science (80-.). (2020). doi:10.1126/science.abd4250

25. Li, Y. et al. Linear epitopes of SARS-CoV-2 spike protein elicit neutralizing antibodies in COVID-19 patients. Cellular and Molecular Immunology 17, 1095–1097 (2020).

26. Kamath, K. et al. Antibody epitope repertoire analysis enables rapid antigen discovery and multiplex serology. Sci. Rep. 10, 1–9 (2020).

27. Pantazes, R. J. et al. Identification of disease-specific motifs in the antibody specificity repertoire via next-generation sequencing. Sci. Rep. 6, (2016).

28. Haynes, W. A., Kamath, K., Daugherty, P. S. & Shon, J. C. Protein-based Immunome Wide Association Studies (PIWAS) for the discovery of significant disease-associated antigens. bioRxiv 2020.03.18.997759 (2020). doi:10.1101/2020.03.18.997759

29. Marklund, E. et al. Serum-IgG responses to SARS-CoV-2 after mild and severe COVID-19 infection and analysis of IgG non-responders. PLoS One 15, e0241104 (2020).

30. Hu, W. T. et al. Antibody Profiles According to Mild or Severe SARS-CoV-2 Infection, Atlanta, Georgia, USA, 2020. Emerg. Infect. Dis. 26, (2020).

31. Benton, D. J. et al. Receptor binding and priming of the spike protein of SARS-CoV-2 for membrane fusion. Nature 1–8 (2020). doi:10.1038/s41586-020-2772-0

32. Wrobel, A. G. et al. SARS-CoV-2 and bat RaTG13 spike glycoprotein structures inform on virus evolution and furin-cleavage effects. Nat. Struct. Mol. Biol. 27, 763–767 (2020).

33. Anand, P., Puranik, A., Aravamudan, M., Venkatakrishnan, A. J. & Soundararajan, V. SARS-CoV-2 strategically mimics proteolytic activation of human ENaC. Elife (2020). doi:10.7554/eLife.58603

34. Hachim, A. et al. ORF8 and ORF3b antibodies are accurate serological markers of early and late SARS-CoV-2 infection. Nat. Immunol. 21, (2020).

35. Young, B. E. et al. Effects of a major deletion in the SARS-CoV-2 genome on the severity of infection and the inflammatory response: an observational cohort study. Lancet 396, 603–611 (2020).

36. Elbe, S. & Buckland-Merrett, G. Data, disease and diplomacy: GISAID’s innovative contribution to global health. Glob. Challenges 1, 33–46 (2017).

37. Shu, Y. & McCauley, J. GISAID: Global initiative on sharing all influenza data – from vision to reality. Eurosurveillance (2017). doi:10.2807/1560-7917.ES.2017.22.13.30494

38. Poh, C. M. et al. Two linear epitopes on the SARS-CoV-2 spike protein that elicit neutralising antibodies in COVID-19 patients. Nat. Commun. (2020). doi:10.1038/s41467-020-16638-2

39. Wang, X. et al. Neutralizing Antibody Responses to Severe Acute Respiratory Syndrome Coronavirus 2 in Coronavirus Disease 2019 Inpatients and Convalescent Patients. Clin. Infect. Dis. (2020). doi:10.1093/cid/ciaa721

40. Zhang, L. et al. Antibody responses against SARS coronavirus are correlated with disease outcome of infected individuals. J. Med. Virol. 78, 1–8 (2006).

41. Schaedel, C. et al. Lung symptoms in pseudohypoaldosteronism type 1 are associated with deficiency of the α-subunit of the epithelial sodium channel. J. Pediatr. 135, 739–745 (1999).

42. Geller, D. S. et al. Autosomal dominant pseudohypoaldosteronism type 1: Mechanisms, evidence for neonatal lethality, and phenotypic expression in adults. J. Am. Soc. Nephrol. 17, 1429–1436 (2006).

43. Gentzsch, M. & Rossier, B. C. A Pathophysiological Model for COVID-19: Critical Importance of Transepithelial Sodium Transport upon Airway Infection. Function 1, (2020).

44. Qiao, B. & De La Cruz, M. O. Enhanced binding of SARS-CoV-2 spike protein to receptor by distal polybasic cleavage sites. ACS Nano 14, 10616–10623 (2020).

45. Zhang, Y. et al. Coagulopathy and Antiphospholipid Antibodies in Patients with Covid-19. N. Engl. J. Med. (2020). doi:10.1056/nejmc2007575

46. Zhang, Y. et al. The ORF8 Protein of SARS-CoV-2 Mediates Immune Evasion through Potently Downregulating MHC-I. bioRxiv (2020). doi:10.1101/2020.05.24.111823

47. Anderson, E. M. et al. Seasonal human coronavirus antibodies are boosted upon SARS-CoV-2 infection but not associated with protection. medRxiv 7, 2020.11.06.20227215 (2020).

48. Ng, K. W. et al. Preexisting and de novo humoral immunity to SARS-CoV-2 in humans. Science (80-.). eabe1107 (2020). doi:10.1126/science.abe1107

49. Poston, D. et al. Absence of SARS-CoV-2 neutralizing activity in pre-pandemic sera from individuals with recent seasonal coronavirus infection. medRxiv (2020). doi:10.1101/2020.10.08.20209650

50. Wang, L. et al. Importance of Neutralizing Monoclonal Antibodies Targeting Multiple Antigenic Sites on the Middle East Respiratory Syndrome Coronavirus Spike Glycoprotein To Avoid Neutralization Escape. J. Virol. (2018). doi:10.1128/jvi.02002-17

51. Sui, J. et al. Effects of Human Anti-Spike Protein Receptor Binding Domain Antibodies on Severe Acute Respiratory Syndrome Coronavirus Neutralization Escape and Fitness. J. Virol. 88, 13769–13780 (2014).

52. Coughlin, M. M. & Prabhakar, B. S. Neutralizing human monoclonal antibodies to severe acute respiratory syndrome coronavirus: target, mechanism of action, and therapeutic potential. Rev. Med. Virol. 22, 2–17 (2012).

53. Hansen, J. et al. Studies in humanized mice and convalescent humans yield a SARS-CoV-2 antibody cocktail. Science (80-.). 369, 1010–1014 (2020).

54. Baum, A. et al. Antibody cocktail to SARS-CoV-2 spike protein prevents rapid mutational escape seen with individual antibodies. Science (80-.). 369, 1014–1018 (2020).

55. Yurkovetskiy, L. et al. SARS-CoV-2 Spike protein variant D614G increases infectivity and retains sensitivity to antibodies that target the receptor binding domain. bioRxiv (2020). doi:10.1101/2020.07.04.187757

56. Walls, A. C. et al. Structure, Function, and Antigenicity of the SARS-CoV-2 Spike Glycoprotein. Cell (2020). doi:10.1016/j.cell.2020.02.058

57. Fernández, A. Structural Impact of Mutation D614G in SARS-CoV-2 Spike Protein: Enhanced Infectivity and Therapeutic Opportunity. ACS Medicinal Chemistry Letters 11, 1667–1670 (2020).

58. Hu, J. et al. The D614G mutation of SARS-CoV-2 spike protein enhances viral infectivity and decreases neutralization sensitivity to individual convalescent sera. bioRxiv (2020).

59. Zhang, L. et al. The D614G mutation in the SARS-CoV-2 spike protein reduces S1 shedding and increases infectivity. bioRxiv Prepr. Serv. Biol. (2020). doi:10.1101/2020.06.12.148726

60. Chen, J., Wang, R., Wang, M. & Wei, G. W. Mutations Strengthened SARS-CoV-2 Infectivity. J. Mol. Biol. 432, 5212–5226 (2020).

61. UniProt: the universal protein knowledgebase. Nucleic Acids Res. 45, D158–D169 (2017).

62. Sanner, M. F., Olson, A. J. & Spehner, J. C. Reduced surface: An efficient way to compute molecular surfaces. Biopolymers 38, 305–320 (1996).

63. Huang, J., Gutteridge, A., Honda, W. & Kanehisa, M. MIMOX: A web tool for phage display based epitope mapping. BMC Bioinformatics (2006). doi:10.1186/1471-2105-7-451

64. Mayrose, I. et al. Pepitope: Epitope mapping from affinity-selected peptides. Bioinformatics (2007). doi:10.1093/bioinformatics/btm493

65. Berman, H. M. et al. The Protein Data Bank. Nucleic Acids Research 28, 235–242 (2000).

66. Burley, S. K. et al. RCSB Protein Data Bank: Biological macromolecular structures enabling research and education in fundamental biology, biomedicine, biotechnology and energy. Nucleic Acids Res. 47, D464–D474 (2019).

67. Hamelryck, T. & Manderick, B. PDB file parser and structure class implemented in Python. Bioinformatics (2003). doi:10.1093/bioinformatics/btg299

68. Cock, P. J. A. et al. Biopython: Freely available Python tools for computational molecular biology and bioinformatics. Bioinformatics 25, 1422–1423 (2009).

69. DeLano, W. L. Pymol: An open-source molecular graphics tool. CCP4 Newsl. Protein Crystallogr. (2002).

70. Tibshirani, R. & Hastie, T. Outlier sums for differential gene expression analysis. Biostatistics 8, 2–8 (2007).

71. Murtagh, F. & Legendre, P. Ward’s Hierarchical Agglomerative Clustering Method: Which Algorithms Implement Ward’s Criterion? J. Classif. (2014). doi:10.1007/s00357-014-9161-z

72. Madeira, F. et al. The EMBL-EBI search and sequence analysis tools APIs in 2019. Nucleic Acids Res. (2019). doi:10.1093/nar/gkz268

