## Supplementary figures and images for "High-resolution mapping and characterization of epitopes in COVID-19 patients"

### Supplemental Figures

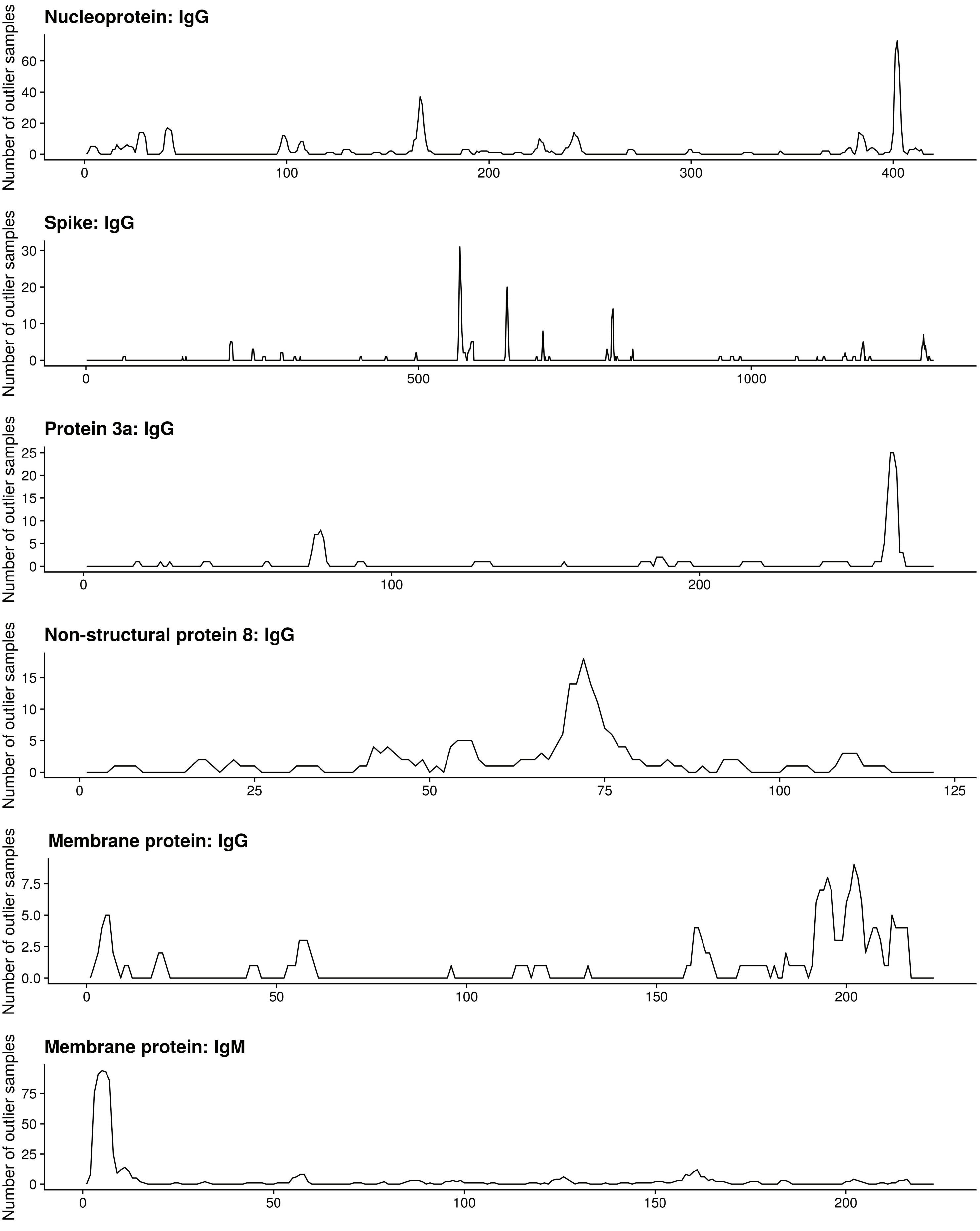

#### Figure S2

YWXYFXK

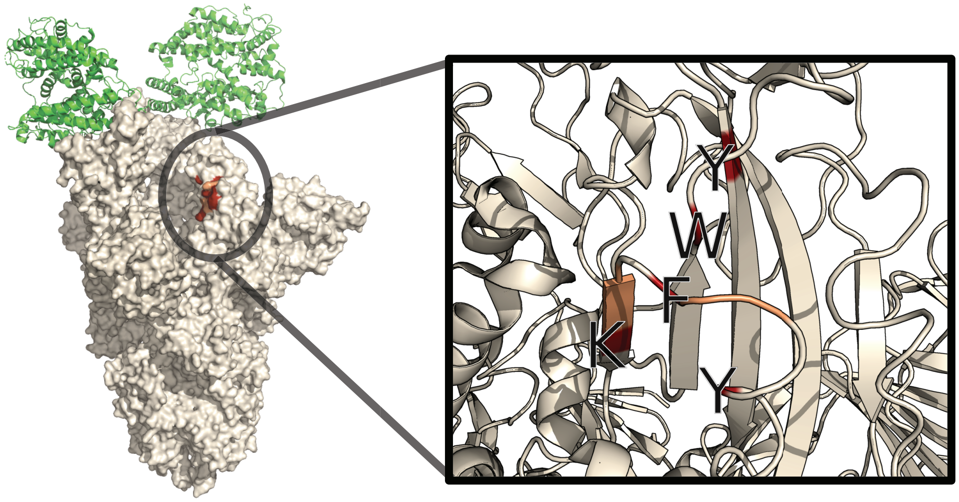


PDB ID: 7A97

YXXYFXK: site #2

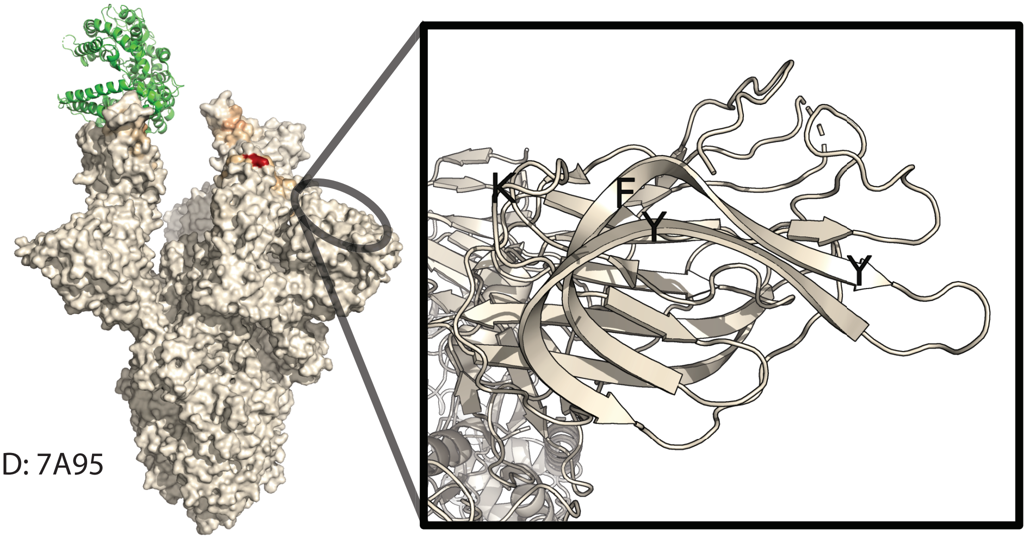


PDB ID: 7A95

YXXYFXK: site #3

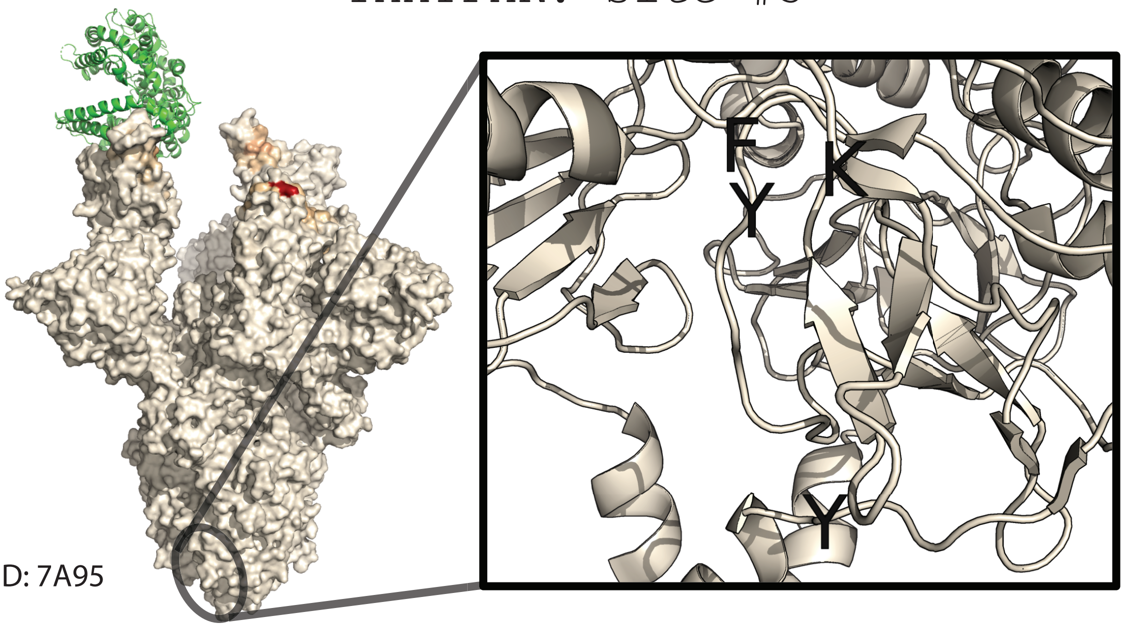


PDB ID: 7A95

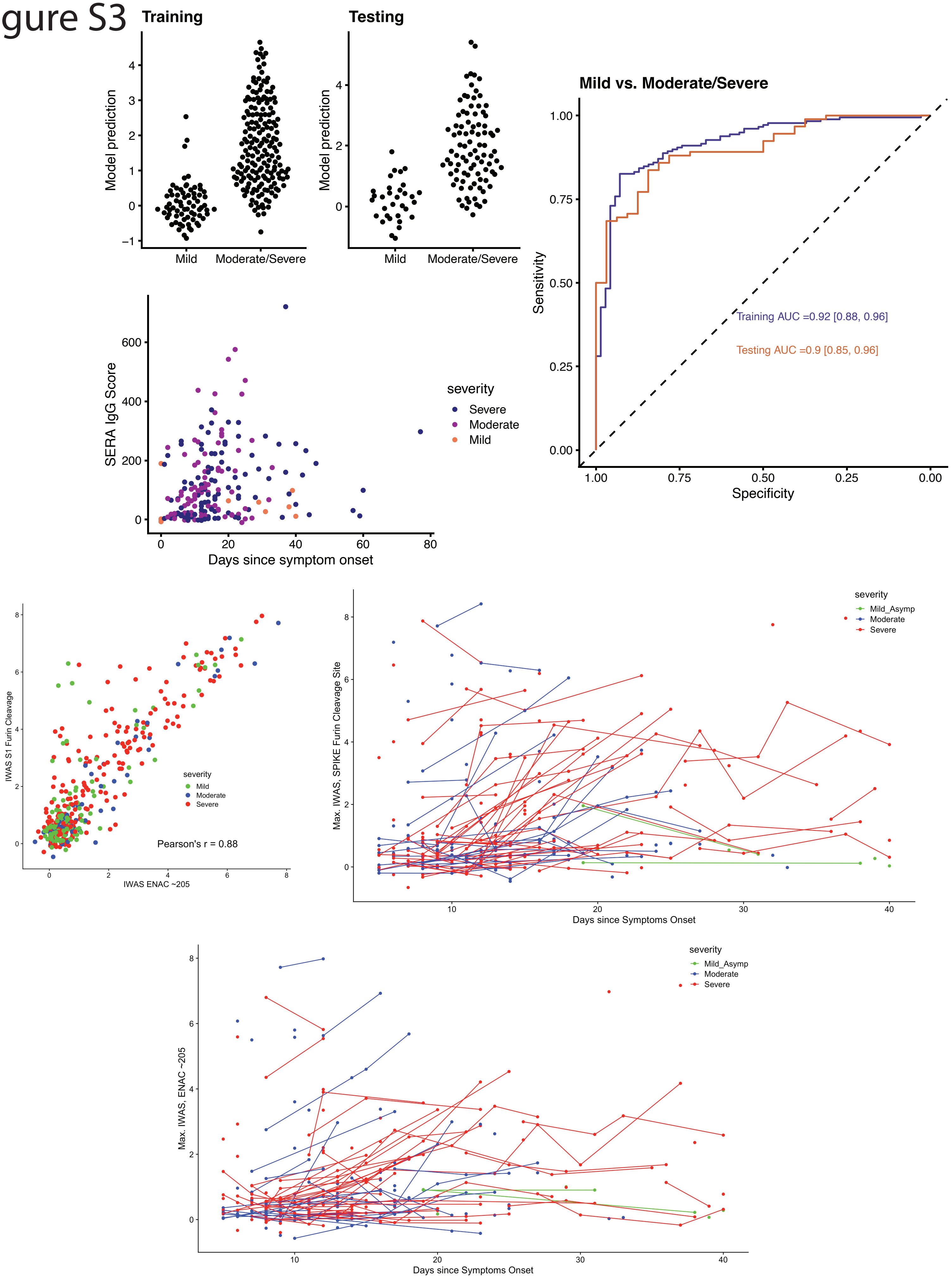

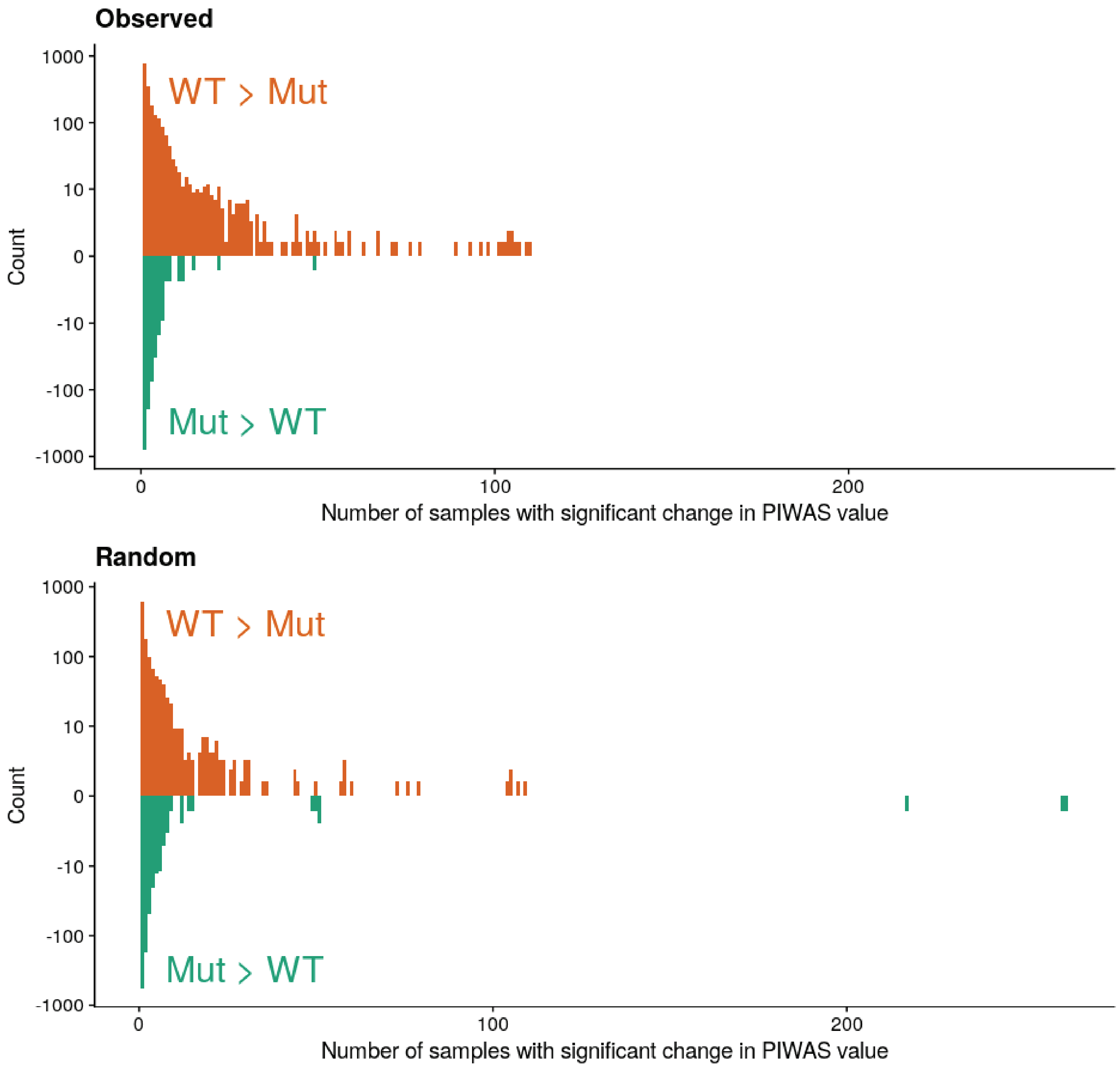
